# NF-κB Mediated Thyroid Inflammation and Its Implications for the Risk of Follicular Thyroid Carcinoma: a systematic review

**DOI:** 10.1101/2024.08.12.24311884

**Authors:** Ovais Shafi, Raveena, Fatima Arshad, Finza Kanwal, Osama Jawed Khan, Muhammad Ashar, Abdul Jabbar, Haresh Kumar, Rahimeen Rajpar, Khizer Sohail, Manwar Madhwani, Muhammad Waqas

**Affiliations:** Sindh Medical College - Jinnah Sindh Medical University / Dow University of Health Sciences, Karachi, Pakistan; Aga Khan University Hospital, Pakistan; SMBBMC, Karachi, Pakistan

**Keywords:** Thyroiditis, Inflammation, Follicular Thyroid Carcinoma, Oncogenesis

## Abstract

**Objective:** The objective of this study is to determine how NF-κB-mediated inflammation disrupts thyroid specific genes/TFs/signaling pathways and the resultant increase in the risk of Follicular Thyroid Carcinoma.

**Background:** Thyroid follicular carcinoma (FTC) is linked to chronic inflammation and NF-κB activation, which disrupts key thyroid-specific genes/transcription factors and signaling pathways. This dysregulation leads to impaired differentiation and increased proliferation, fostering oncogenesis. The interplay between NF-κB and thyroid components is critical for developing possible targeted therapies. This study aims to focus on these mechanisms, with the future potential of possibly improving diagnostic, prognostic, and therapeutic strategies for FTC.

**Methods:** Databases, including PubMed, MEDLINE, Google Scholar, and open access/ subscription-based journals were searched for published articles without any date restrictions, to investigate how NF-κB-mediated inflammation disrupts thyroid specific genes/TFs/signaling pathways and the resultant increase in the risk of Follicular Thyroid Carcinoma. Based on the criteria mentioned in the methods section, studies were systematically reviewed to investigate the research question. This study adheres to relevant PRISMA guidelines (Preferred Reporting Items for Systematic Reviews and Meta-Analyses).

**Results:** NF-κB activation disrupts key thyroid transcription factors and signaling pathways, leading to increased risk of thyroid follicular carcinoma. Dysregulation of NKX2-1, PAX8, FOXE1, and HHEX impairs thyroid development and differentiation. NF-κB alters TSH signaling and disrupts RET/PTC, leading to aberrant cell proliferation. It further impacts the MAPK/ERK, PI3K/AKT, and Wnt/β- catenin pathways, driving oncogenic transformations. Additionally, NF-κB-induced changes in FGF, Shh, BMP, Notch, and TGF beta pathways exacerbate tumorigenesis by promoting abnormal cell growth and survival, collectively increasing follicular carcinoma risk.

**Conclusion:** Chronic inflammation and NF-κB activation disrupt key transcription factors (NKX2-1, PAX8, FOXE1, HHEX) and signaling pathways (TSH, RET/PTC, MAPK/ERK, PI3K/AKT, Wnt/β-catenin, FGF, Shh, BMP, Notch, TGF-β) essential for thyroid development and function. This dysregulation leads to impaired differentiation, increased proliferation, and enhanced survival of thyroid follicular cells, fostering genomic instability and oncogenic transformation. These findings highlight the role of NF-κB in promoting follicular thyroid carcinoma (FTC) and points towards the need for targeted therapies to restore normal signaling and prevent malignancy in thyroid diseases.

## Background

Thyroid follicular carcinoma (FTC) represents a significant clinical challenge due to its propensity for late diagnosis and potential for metastasis. Recent studies have highlighted the key role of chronic inflammation in the pathogenesis of various cancers, including FTC. Central to the inflammatory response is the nuclear factor kappa-light-chain-enhancer of activated B cells (NF-κB) signaling pathway, which regulates the expression of genes involved in immune and inflammatory responses [1]. Chronic activation of NF-κB has been implicated in the dysregulation of multiple cellular processes, including proliferation, differentiation, and survival, which are critical in maintaining thyroid gland homeostasis [2]. Thyroid follicular cells rely on a delicate balance of specific transcription factors (TFs) such as NKX2-1, PAX8, FOXE1, and HHEX, and signaling pathways like TSH, RET/PTC, MAPK/ERK, PI3K/AKT, Wnt/β-catenin, FGF, Shh, BMP, Notch, and TGF-β to ensure proper development, differentiation, and function [3]. Dysregulation of these TFs and pathways disrupts thyroid follicular cell fate, leading to dedifferentiation and increased proliferation, hallmarks of oncogenic transformation. However, the precise mechanisms by which NF-κB-induced inflammation contributes to these dysregulations and predisposes the thyroid to FTC remain poorly understood. The need for this research study arises from the critical gap in understanding the molecular interplay between NF-κB signaling and thyroid-specific genes and pathways [4], [5]. Focusing on these mechanisms is essential for developing possible targeted therapies aimed at preventing and treating FTC. By comprehensively investigating how NF-κB damages these molecular components, this study aims to provide new insights into the pathogenesis of FTC, ultimately contributing to improved diagnostic, prognostic, and therapeutic strategies. The findings could pave the way for possible new interventions that mitigate inflammation-induced thyroid carcinogenesis, thereby enhancing patient outcomes and reducing the burden of this malignancy [6, 7, 8].

## Methods

### Aim of the Study

The aim of this study is to investigate how NF-κB damages thyroid genes, transcription factors, and signaling pathways (NKX2-1, PAX8, FOXE1 (TTF-2), HHEX, TSH, RET/PTC, MAPK/ERK Pathway, PI3K/AKT Pathway, Wnt/β-catenin Pathway, FGF, Shh, BMP, Notch, TGF beta) and the resultant increase in the risk of Thyroid Follicular Carcinoma. This includes examining the impact of NF-κB dysregulation on these factors and understanding the mechanisms by which inflammation contributes to thyroid tumorigenesis. These are also the limitations of the study.

### Research Question

How does NF-κB-mediated inflammation disrupt key thyroid genes, transcription factors, and signaling pathways, leading to an increased risk of Thyroid Follicular Carcinoma?

### Search Focus

A comprehensive literature search was conducted using the PUBMED database, MEDLINE database, and Google Scholar, as well as open access and subscription-based journals. There were no date restrictions for published articles. The search focused on the following key genes, transcription factors, and signaling pathways involved in thyroid function and inflammation, specifically investigating their roles in the context of NF-κB-mediated damage and thyroid follicular carcinoma risk:

- NKX2-1
- PAX8
- FOXE1 (TTF-2)
- HHEX
- TSH (Thyroid Stimulating Hormone)
- RET/PTC (Rearranged during Transfection/ Papillary Thyroid Carcinoma)
- MAPK/ERK Pathway (Mitogen-Activated Protein Kinase/ Extracellular Signal-Regulated Kinase)
- PI3K/AKT Pathway (Phosphoinositide 3-Kinase/ Protein Kinase B)
- Wnt/β-catenin Pathway
- FGF (Fibroblast Growth Factor)
- Shh (Sonic Hedgehog)
- BMP (Bone Morphogenetic Protein)
- Notch
- TGF beta (Transforming Growth Factor beta)

Screening of the literature was also done on this same basis and related data was extracted. Literature search began in November 2020 and ended in January 2024. An in-depth investigation was conducted during this duration based on the parameters of the study as defined above. During revision, further literature was searched and referenced until July 2024. The literature search and all sections of the manuscript were checked multiple times during the months of revision (February 2024 – July 2024) to maintain the highest accuracy possible. This comprehensive approach ensured that the selected studies provided valuable insights into the inflammatory pathways and their implications in thyroid follicular carcinoma risk. This study adheres to relevant PRISMA guidelines (Preferred Reporting Items for Systematic Reviews and Meta-Analyses).

### Search Queries/Keywords

1. **General Terms:**

“Thyroid Follicular Carcinoma” “Thyroid cancer”
“NF-κB and thyroid”
“Thyroid Genes/TFs/Signaling Pathways”
2. **Specific Genes and Pathways:**

“NF-κB” AND “NKX2-1” AND “Thyroid Follicular Carcinoma”
“NF-κB” AND “PAX8” AND “Thyroid Follicular Carcinoma”
“NF-κB” AND “FOXE1” AND “Thyroid Follicular Carcinoma”
“NF-κB” AND “HHEX” AND “Thyroid Follicular Carcinoma”
“NF-κB” AND “TSH” AND “Thyroid Follicular Carcinoma”
“NF-κB” AND “RET/PTC” AND “Thyroid Follicular Carcinoma”
“NF-κB” AND “MAPK/ERK Pathway” AND “Thyroid Follicular Carcinoma”
“NF-κB” AND “PI3K/AKT Pathway” AND “Thyroid Follicular Carcinoma”
“NF-κB” AND “Wnt/β-catenin Pathway” AND “Thyroid Follicular Carcinoma”
“NF-κB” AND “FGF” AND “Thyroid Follicular Carcinoma”
“NF-κB” AND “Shh” AND “Thyroid Follicular Carcinoma”
“NF-κB” AND “BMP” AND “Thyroid Follicular Carcinoma”
“NF-κB” AND “Notch” AND “Thyroid Follicular Carcinoma”
“NF-κB” AND “TGF beta” AND “Thyroid Follicular Carcinoma”

Boolean operators (AND, OR) were utilized to construct search queries in relation to the following terms: “Thyroid Follicular Carcinoma,” “NF-κB,” “Inflammation,” and “Thyroid Inflammation.” These operators were applied in relation to key genes, transcription factors, and signaling pathways involved in thyroid function. This approach facilitated the comprehensive retrieval of relevant articles by combining these terms to capture studies focusing on their interrelations and individual contributions to thyroid tumorigenesis.

### Objectives of the Searches

- To determine the impact of NF-κB-mediated inflammation on disrupting thyroid genes, transcription factors, and signaling pathways.
- To investigate how such disruptions contribute to thyroid tumorigenesis.
- To assess the role of specific genes, TFs and pathways (NKX2-1, PAX8, FOXE1, HHEX, TSH, RET/PTC, MAPK/ERK Pathway, PI3K/AKT Pathway, Wnt/β-catenin Pathway, FGF, Shh, BMP, Notch, TGF beta) in the increased risk of thyroid follicular carcinoma.

### Screening and Eligibility Criteria

#### Initial Screening

Articles were initially screened based on titles and abstracts to identify direct relevance to the study objectives.

#### Full-Text Review

Articles that passed the initial screening underwent a comprehensive full-text review. Articles were included if they provided valuable insights into the roles of the specified genes, transcription factors, and signaling pathways in thyroid follicular carcinoma in the context of NF-κB-mediated inflammation.

#### Data Extraction

Relevant data was extracted from each selected article, focusing on key findings and outcomes related to the study objectives.

### Inclusion and Exclusion Criteria

#### Inclusion Criteria

- Articles directly related to the key genes, transcription factors, and signaling pathways (NKX2-1, PAX8, FOXE1, HHEX, TSH, RET/PTC, MAPK/ERK Pathway, PI3K/AKT Pathway, Wnt/β-catenin Pathway, FGF, Shh, BMP, Notch, TGF beta) involved in thyroid development, function and inflammation.
- Studies focusing on the impact of NF-κB dysregulation on these factors and thyroid tumorigenesis.

### Exclusion Criteria

- Articles that did not conform to the study focus.
- Insufficient methodological rigor.
- Data not aligning with the research questions.

### Rationale for Screening and Inclusion

- **NKX2-1:** Essential for thyroid development and function. Disruption leads to tumorigenesis.
- **PAX8:** Regulates thyroid-specific gene expression. Dysregulation associated with thyroid cancer.
- **FOXE1 (TTF-2):** Involved in thyroid development. Aberrant expression linked to cancer.
- **HHEX:** Plays a role in thyroid organogenesis. Dysregulation contributes to cancer.
- **TSH:** Stimulates thyroid hormone production. Dysregulated signaling linked to tumor growth.
- **RET/PTC:** Oncogene involved in thyroid carcinogenesis. Abnormal expression leads to cancer.
- **MAPK/ERK Pathway:** Key pathway in cell proliferation and differentiation. Dysregulation promotes cancer cell growth.
- **PI3K/AKT Pathway:** Critical for cell survival and growth. Aberrant signaling linked to cancer.
- **Wnt/β-catenin Pathway:** Regulates cell fate and proliferation. Dysregulation associated with cancer.
- **FGF:** Important for cell growth and differentiation. Dysregulation contributes to cancer development.
- **Shh:** Involved in thyroid organogenesis. Aberrant signaling linked to tumor growth.
- **BMP:** Regulates tissue homeostasis. Abnormal signaling linked to cancer.
- **Notch:** Controls cell differentiation and proliferation. Dysregulation leads to cancer.
- **TGF beta:** Regulates cell growth and apoptosis. Dysregulation promotes tumor progression.

This comprehensive screening and inclusion rationale ensure that the selected studies provide valuable insights into the roles of these critical genes, transcription factors, and signaling pathways in thyroid inflammation and tumorigenesis. PRISMA Flow Diagram is Fig 1.

**Fig 1.**
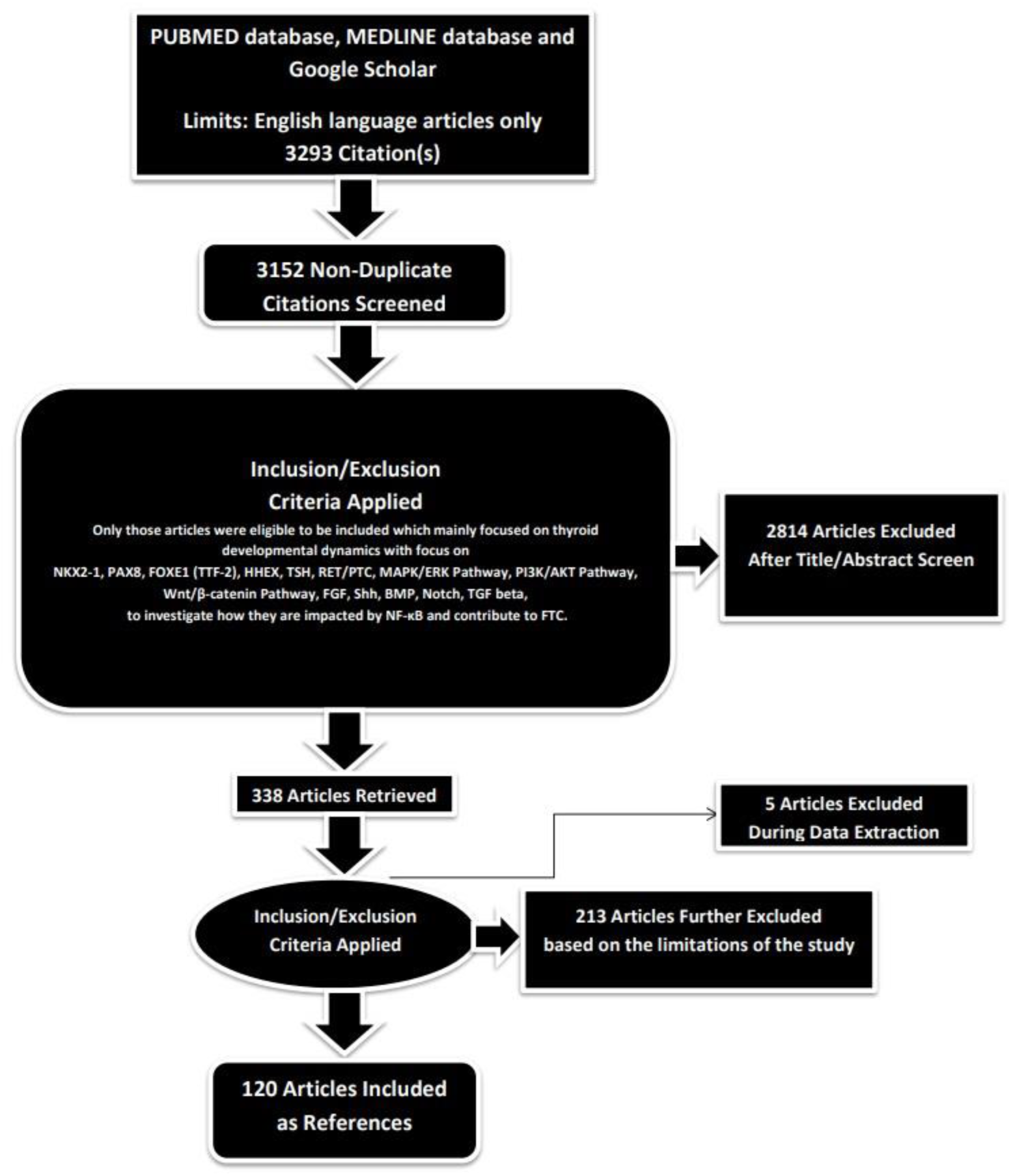
PRISMA FLOW DIAGRAM. This figure represents graphically the flow of citations in the study.

### Assessment of Article Quality and Potential Biases

Ensuring the quality and minimizing potential biases of the selected articles were crucial aspects to guarantee the rigor and reliability of the research findings.

#### Quality Assessment

The initial step in quality assessment involved evaluating the methodological rigor of the selected articles. This included a thorough examination of the study design, data collection methods, and analyses conducted. The significance of the study’s findings was weighed based on the quality of the evidence presented. Articles demonstrating sound methodology— such as well-designed studies, controlled variables, and scientifically robust data—were considered of higher quality. Peer-reviewed articles, scrutinized by experts in the field, served as a significant indicator of quality.

### Potential Biases Assessment

- **Publication Bias:** To address the potential for publication bias, a comprehensive search strategy was adopted to include a balanced representation of both positive and negative results, incorporating a wide range of published articles from databases like Google Scholar.
- **Selection Bias:** Predefined and transparent inclusion criteria were applied to minimize subjectivity in the selection process. Articles were chosen based on their relevance to the study’s objectives, adhering strictly to these criteria. This approach reduced the risk of subjectivity and ensured that the selection process was objective and consistent.
- **Reporting Bias:** To mitigate reporting bias, articles were checked for inconsistencies or missing data. Multiple detailed reviews of the methodologies and results were conducted for all selected articles to identify and address any reporting bias.

By including high-quality, peer-reviewed studies and thoroughly assessing potential biases, this study aimed to provide a robust foundation for the results and conclusions presented.

### Language and Publication Restrictions

We restricted our selection to publications in the English language. There were no limitations imposed on the date of publication. Unpublished studies were not included in our analysis.

## Results

A total of 3293 articles were identified using database searching, and 3152 were recorded after duplicates removal. 2814 were excluded after screening of title/abstract, 213 were finally excluded, and 5 articles were excluded during data extraction. These exclusions were primarily due to factors such as non-conformity with the study focus, insufficient methodological rigor, or data that did not align with our research questions. Finally, 120 articles were included as references.

### Investigating Thyroid Genes/TFs/Signaling Pathways

#### 1. NKX2-1

##### Role of NF-κB in disrupting it

NKX2-1, also known as thyroid transcription factor-1 (TTF-1), is essential for the development and function of the thyroid gland. Dysregulation of NKX2-1 due to inflammation, including the involvement of NF-κB (nuclear factor kappa-light-chain-enhancer of activated B cells), can have significant implications for thyroid function and health. Inflammatory cytokines such as TNF-α (tumor necrosis factor-alpha), IL-1β (interleukin-1 beta), and IL-6 are often elevated during thyroid inflammation (thyroiditis) [9]. These cytokines can influence the expression and activity of transcription factors, including NKX2-1, through signaling pathways that modulate gene transcription. NF-κB is a key transcription factor activated during inflammation, regulating the expression of various genes involved in immune and inflammatory responses. Activation of NF-κB can lead to increased production of inflammatory cytokines, creating a feedback loop that sustains inflammation. NF-κB can also directly interfere with the expression and function of NKX2-1 by binding to the promoter regions of genes regulated by NKX2-1 or by competing for coactivators [10].

Inflammation often results in increased oxidative stress within thyroid cells, which can modify transcription factor activity through post-translational modifications (e.g., phosphorylation, acetylation), potentially altering NKX2-1 function and stability. Inflammatory processes can induce epigenetic changes, such as DNA methylation and histone modifications, which can affect gene expression. Inflammatory cytokines and NF-κB activation can lead to changes in the epigenetic landscape of thyroid cells, potentially silencing or reducing the expression of NKX2-1. NKX2-1 is a master regulator of thyroid-specific genes, including thyroglobulin (TG), thyroperoxidase (TPO), and the sodium/iodide symporter (NIS). Inflammation and NF-κB activation can disrupt the expression of these genes, either directly or through the downregulation of NKX2-1, impairing thyroid hormone synthesis and secretion. Inflammatory signaling pathways often interact with other cellular signaling pathways, such as Wnt, MAPK, and PI3K/AKT. Cross-talk between NF-κB and these pathways can modulate the activity of NKX2-1, either through direct interactions or by altering the cellular context in which NKX2-1 operates. Chronic inflammation often involves the infiltration of immune cells, such as macrophages and lymphocytes, into the thyroid tissue [11]. These immune cells can produce inflammatory cytokines and other mediators that impact thyroid follicular cells, leading to altered expression and function of NKX2-1. There are several potential mechanisms of NKX2-1 dysregulation in thyroid inflammation. NF-κB may directly bind to NKX2-1 promoter regions or compete for coactivators, leading to decreased NKX2-1 expression. Inflammatory cytokines may induce DNA methylation or histone modifications in the NKX2-1 gene locus, reducing its expression. Inflammatory mediators and oxidative stress may alter NKX2-1 through phosphorylation, acetylation, or other modifications, affecting its stability and function. Chronic inflammation may disrupt the signaling networks that normally support NKX2-1 function, such as TSH signaling, leading to impaired thyroid cell differentiation and hormone production [12].

##### Resultant dysregulation in thyroid cell fate and resultant impact on FTC development

The impact of NF-κB on NKX2-1 can disrupt thyroid follicular cell fate and predispose the thyroid gland to follicular thyroid carcinoma (FTC) through several mechanisms. Chronic inflammation in the thyroid gland, often caused by autoimmune thyroiditis or other inflammatory conditions, leads to the activation of NF-κB. NF-κB is activated by pro-inflammatory cytokines (e.g., TNF-α, IL-1β) and through signaling pathways involving the IκB kinase (IKK) complex, which phosphorylates IκB proteins, leading to their degradation and the release of NF-κB dimers that translocate to the nucleus [13]. NF-κB can directly bind to the promoter region of the NKX2-1 gene or to co-regulators, inhibiting its transcription. Inflammatory cytokines and NF-κB can induce epigenetic modifications (e.g., DNA methylation, histone modifications) that silence or reduce NKX2-1 expression. NKX2-1 is a critical transcription factor for thyroid follicular cell differentiation and function. It regulates the expression of thyroid-specific genes, including thyroglobulin (TG), thyroperoxidase (TPO), and the sodium/iodide symporter (NIS). NKX2-1 ensures proper differentiation of thyroid follicular cells and maintenance of their functional phenotype, including hormone production. Inhibition of NKX2-1 by NF-κB leads to reduced expression of key thyroid-specific genes, impairing the differentiation and function of follicular cells [14]. The downregulation of NKX2-1 can cause thyroid follicular cells to lose their differentiated state and adopt a more undifferentiated or precursor-like phenotype. NF-κB activation promotes cell survival and proliferation by inducing anti-apoptotic genes and pro-proliferative signals, contributing to the survival of dysregulated cells. The loss of NKX2-1 and the accompanying dysregulation of thyroid-specific gene expression can be an early event in the pathogenesis of FTC. Chronic NF-κB activation supports uncontrolled cell proliferation and resistance to apoptosis, creating a pro-tumorigenic environment. Inflammatory conditions and oxidative stress associated with chronic NF-κB activation can lead to DNA damage and genomic instability, further contributing to cancer development. NKX2-1 inhibition leads to the dedifferentiation of thyroid follicular cells, a hallmark of cancer development. NF-κB promotes the expression of genes that enhance cell proliferation and survival, providing a growth advantage to dysregulated cells. NF-κB activation results in the upregulation of anti-apoptotic proteins (e.g., Bcl-2, Bcl-xL), helping pre-malignant cells evade programmed cell death [15]. Persistent inflammation mediated by NF-κB creates a microenvironment that favors carcinogenesis, including increased angiogenesis and immune evasion. The impact of NF-κB on NKX2-1 disrupts thyroid follicular cell fate by downregulating key thyroid-specific genes and promoting dedifferentiation. This disruption, combined with the pro-survival and proliferative signals from NF-κB, predisposes the thyroid gland to follicular thyroid carcinoma (FTC). The chronic inflammatory environment and associated oxidative stress further contribute to genomic instability and tumorigenesis, making the NF-κB/NKX2-1 interaction a critical factor in the development of FTC [16].

#### 2. PAX8

##### Role of NF-κB in disrupting it

PAX8 is a essential transcription factor involved in thyroid development and function. Dysregulation of PAX8 due to inflammation, including the involvement of NF-κB, can significantly impact thyroid health and predispose the thyroid to various pathologies [17]. Pro-inflammatory cytokines such as TNF-α, IL-1β, and IL-6 are commonly elevated in thyroid inflammation (e.g., thyroiditis). These cytokines can modulate the expression and activity of transcription factors, including PAX8, by activating signaling pathways that alter gene transcription and protein function. NF-κB is a key transcription factor in inflammatory responses, regulating genes involved in immune and inflammatory responses. Activation of NF-κB can influence PAX8 expression and function through several mechanisms. NF-κB may compete with or inhibit transcription factors and coactivators that regulate PAX8 expression. Additionally, NF-κB activation can lead to changes in the epigenetic landscape, such as DNA methylation and histone modifications, that suppress PAX8 expression [18]. Continuous NF-κB activation maintains a pro-inflammatory state that further dysregulates PAX8. Inflammation often leads to increased oxidative stress in thyroid cells. Oxidative stress can modify transcription factors, including PAX8, through post-translational modifications (e.g., phosphorylation, acetylation), affecting their stability and function. PAX8 is essential for the expression of thyroid-specific genes, such as thyroglobulin (TG), thyroperoxidase (TPO), and the sodium/iodide symporter (NIS). Inflammatory cytokines and NF-κB can disrupt the expression of these genes, either directly or through downregulation of PAX8, impairing thyroid hormone synthesis and secretion. Inflammatory signaling pathways interact with other cellular pathways, such as Wnt, MAPK, and PI3K/AKT [19]. Cross-talk between NF-κB and these pathways can alter the activity of PAX8, either through direct interactions or by changing the cellular context in which PAX8 operates. Chronic inflammation involves the infiltration of immune cells, such as macrophages and lymphocytes, into the thyroid tissue. These immune cells produce inflammatory cytokines and other mediators that impact thyroid follicular cells, leading to altered expression and function of PAX8. There are several potential mechanisms of PAX8 dysregulation in thyroid inflammation. NF-κB may compete with transcription factors that regulate PAX8 or inhibit its expression by binding to co-regulators.

Inflammatory cytokines and NF-κB can induce DNA methylation or histone modifications in the PAX8 gene locus, reducing its expression. Inflammatory mediators and oxidative stress can modify PAX8 through phosphorylation, acetylation, or other modifications, affecting its stability and function. Chronic inflammation can disrupt signaling networks that support PAX8 function, such as TSH signaling, leading to impaired thyroid cell differentiation and hormone production. Inflammation, particularly through the activation of NF-κB, can dysregulate PAX8 in multiple ways, including direct inhibition, epigenetic silencing, and post-translational modifications. This dysregulation can disrupt thyroid follicular cell fate by impairing the expression of thyroid-specific genes and leading to dedifferentiation. The chronic inflammatory environment and associated oxidative stress can further contribute to genomic instability and thyroid pathologies, including thyroid cancer [20].

##### Resultant dysregulation in thyroid cell fate and resultant impact on FTC development

The impact of NF-κB on PAX8 can disrupt thyroid follicular cell fate and predispose the thyroid gland to follicular thyroid carcinoma (FTC) through several mechanisms. Chronic inflammation in the thyroid gland, such as that seen in autoimmune thyroiditis or other inflammatory conditions, leads to the activation of NF-κB. NF-κB is activated by pro-inflammatory cytokines (e.g., TNF-α, IL-1β) and through signaling pathways involving the IκB kinase (IKK) complex, which phosphorylates IκB proteins, leading to their degradation and the release of NF-κB dimers that translocate to the nucleus [21]. NF-κB can directly inhibit PAX8 transcription by competing with transcription factors and coactivators essential for PAX8 expression. Additionally, NF-κB activation can induce epigenetic modifications (e.g., DNA methylation, histone modifications) that silence or reduce PAX8 expression. PAX8 is a critical transcription factor for thyroid follicular cell differentiation and function. It regulates the expression of thyroid-specific genes, including thyroglobulin (TG), thyroperoxidase (TPO), and the sodium/iodide symporter (NIS). PAX8 ensures proper differentiation of thyroid follicular cells and maintenance of their functional phenotype, including hormone production [22]. Inhibition of PAX8 by NF-κB leads to reduced expression of key thyroid-specific genes, impairing the differentiation and function of follicular cells. The downregulation of PAX8 can cause thyroid follicular cells to lose their differentiated state and adopt a more undifferentiated or precursor-like phenotype. NF-κB activation promotes cell survival and proliferation by inducing anti-apoptotic genes and pro-proliferative signals, contributing to the survival of dysregulated cells. The loss of PAX8 and the accompanying dysregulation of thyroid-specific gene expression can be an early event in the pathogenesis of FTC. Chronic NF-κB activation supports uncontrolled cell proliferation and resistance to apoptosis, creating a pro-tumorigenic environment. Inflammatory conditions and oxidative stress associated with chronic NF-κB activation can lead to DNA damage and genomic instability, further contributing to cancer development [23]. PAX8 inhibition leads to the dedifferentiation of thyroid follicular cells, a hallmark of cancer development. NF-κB promotes the expression of genes that enhance cell proliferation and survival, providing a growth advantage to dysregulated cells. NF-κB activation results in the upregulation of anti-apoptotic proteins (e.g., Bcl-2, Bcl-xL), helping pre-malignant cells evade programmed cell death. Persistent inflammation mediated by NF-κB creates a microenvironment that favors carcinogenesis, including increased angiogenesis and immune evasion. The impact of NF-κB on PAX8 disrupts thyroid follicular cell fate by downregulating key thyroid-specific genes and promoting dedifferentiation. This disruption, combined with the pro-survival and proliferative signals from NF-κB, predisposes the thyroid gland to follicular thyroid carcinoma (FTC). The chronic inflammatory environment and associated oxidative stress further contribute to genomic instability and tumorigenesis, making the NF-κB/PAX8 interaction a critical factor in the development of FTC [24].

#### 3. FOXE1 (TTF-2)

##### Role of NF-κB in disrupting it

FOXE1 (also known as TTF-2) plays a critical role in thyroid development and function, particularly in the differentiation and maintenance of thyroid follicular cells. Dysregulation of FOXE1 due to inflammation, including the involvement of NF-κB, can have significant implications for thyroid health and disease. Pro-inflammatory cytokines (e.g., TNF-α, IL-1β, IL-6) are elevated during thyroid inflammation and can influence transcription factors like FOXE1 through signaling pathways that alter gene expression and protein function. NF-κB, a key mediator of inflammation, regulates genes involved in immune responses and can impact FOXE1 in several ways [25]. NF-κB may directly inhibit FOXE1 by binding to its promoter regions or competing with transcriptional activators/co-activators. Activation of NF-κB can also lead to epigenetic changes (e.g., DNA methylation, histone modifications) that silence or reduce FOXE1 expression. Additionally, NF-κB activation may induce the expression of microRNAs or other factors that target FOXE1 mRNA stability or translation. Inflammation often leads to increased oxidative stress within thyroid cells, which can modify FOXE1 function through post-translational modifications (e.g., phosphorylation, acetylation), affecting its stability and activity [26]. FOXE1 regulates the expression of key thyroid-specific genes such as thyroglobulin (TG), thyroperoxidase (TPO), and the sodium/iodide symporter (NIS). Inflammatory cytokines and NF-κB activation can disrupt the expression of these genes, either directly or through downregulation of FOXE1, impairing thyroid hormone synthesis and secretion. Inflammatory signaling pathways (e.g., MAPK, PI3K/AKT) can interact with NF-κB and influence FOXE1 activity by modulating its phosphorylation status or intracellular localization. Chronic inflammation involves infiltration of immune cells (e.g., macrophages, lymphocytes) into thyroid tissue, and these immune cells secrete cytokines and other mediators that affect FOXE1 expression and thyroid follicular cell function. Potential mechanisms of FOXE1 dysregulation in thyroid inflammation include direct inhibition by NF-κB, where NF-κB activation inhibits FOXE1 transcription by competing for binding sites or by inducing repressive chromatin modifications [27]. Inflammatory cytokines and oxidative stress can induce DNA methylation and histone modifications in the FOXE1 gene promoter, reducing its expression. Oxidative stress can lead to FOXE1 protein modifications that affect its stability and activity in regulating thyroid-specific genes. Inflammation disrupts signaling pathways necessary for FOXE1 function, impairing its ability to regulate thyroid cell differentiation and hormone production. Inflammation, particularly through NF-κB activation, can dysregulate FOXE1 in multiple ways, leading to disruption of thyroid follicular cell fate and potentially predisposing the thyroid gland to diseases such as thyroiditis and thyroid cancer [28].

##### Resultant dysregulation in thyroid cell fate and resultant impact on FTC development

The impact of NF-κB on FOXE1 (TTF-2) can disrupt thyroid follicular cell fate and predispose the thyroid gland to follicular thyroid carcinoma (FTC) through various mechanisms. Chronic inflammation in the thyroid, such as autoimmune thyroiditis or chronic infection, leads to the activation of NF-κB. This activation is initiated by pro-inflammatory cytokines (e.g., TNF-α, IL-1β) and involves the degradation of IκB proteins, allowing NF-κB dimers to translocate into the nucleus and regulate gene expression [29]. NF-κB can directly inhibit FOXE1 transcription by binding to regulatory regions in the FOXE1 gene or by competing with transcriptional activators/co-activators required for FOXE1 expression. Additionally, NF-κB activation can induce epigenetic changes, such as DNA methylation and histone modifications, in the FOXE1 gene promoter region, leading to reduced FOXE1 expression. NF-κB activation may also stimulate the expression of microRNAs or other factors that target FOXE1 mRNA stability or translation, further reducing FOXE1 levels [30]. FOXE1 plays a key role in thyroid follicular cell differentiation and function by regulating the expression of thyroid-specific genes, including thyroglobulin (TG), thyroperoxidase (TPO), and the sodium/iodide symporter (NIS). Proper FOXE1 activity is essential for maintaining the differentiated state of thyroid follicular cells and ensuring the synthesis and secretion of thyroid hormones. However, inhibition of FOXE1 by NF-κB results in decreased expression of key thyroid-specific genes, impairing thyroid hormone production and follicular cell differentiation. Reduced FOXE1 levels can lead to dedifferentiation of thyroid follicular cells, promoting a more progenitor-like phenotype that is susceptible to oncogenic transformation. NF-κB activation enhances cell survival and proliferation through the upregulation of anti-apoptotic genes and pro-survival pathways, contributing to the expansion of pre-malignant cells. Early dysregulation of FOXE1 and subsequent disruption of thyroid-specific gene expression can initiate the pathogenesis of FTC. NF-κB-driven signaling pathways promote uncontrolled cell proliferation and inhibit apoptosis, providing a growth advantage to pre-malignant thyroid cells. Chronic inflammation and oxidative stress associated with NF-κB activation can induce DNA damage and genomic instability, contributing to the acquisition of malignant traits [31]. FOXE1 inhibition leads to the loss of thyroid follicular cell differentiation, a hallmark of thyroid cancer progression. NF-κB activation sustains proliferative signaling pathways, promoting the expansion of cells with oncogenic potential. Furthermore, NF-κB upregulates anti-apoptotic proteins, protecting dysregulated thyroid cells from programmed cell death. Chronic inflammation mediated by NF-κB creates a tumor-promoting microenvironment characterized by increased angiogenesis, immune evasion, and altered metabolism. NF-κB-mediated disruption of FOXE1 function disrupts thyroid follicular cell fate by impairing the expression of thyroid-specific genes and promoting dedifferentiation. This dysregulation, coupled with NF-κB’s pro-survival and proliferative signals, predisposes the thyroid gland to follicular thyroid carcinoma (FTC) [32].

#### 4. HHEX

##### Role of NF-κB in disrupting it

HHEX (Hematopoietically-expressed homeobox protein) is involved in various developmental processes, including thyroid development, where it plays a key role in regulating thyroid morphogenesis and function. Dysregulation of HHEX due to inflammation, including the involvement of NF-κB, can impact thyroid health and contribute to thyroid disorders. Pro-inflammatory cytokines such as TNF-α, IL-1β, and IL-6 are elevated during thyroid inflammation. These cytokines can modulate HHEX expression and function through signaling pathways that alter gene transcription and protein stability [33]. NF-κB is a central mediator of inflammation and regulates genes involved in immune responses.

NF-κB activation can influence HHEX in several ways: it may directly bind to the regulatory regions of the HHEX gene, leading to transcriptional repression; it can induce the expression of microRNAs or other factors that target HHEX mRNA stability or translation, thereby reducing HHEX levels; and NF-κB-mediated changes in chromatin structure, such as histone modifications and DNA methylation, can alter HHEX expression patterns [34]. Inflammation often leads to increased oxidative stress within thyroid cells. Oxidative stress can modify HHEX function through post-translational modifications, such as phosphorylation and acetylation, affecting its activity and interaction with co-regulatory proteins. HHEX regulates the expression of genes involved in thyroid development and function, including genes encoding transcription factors and signaling molecules critical for thyroid morphogenesis. Inflammatory cytokines and NF-κB activation can disrupt the expression of these genes, either directly through HHEX or indirectly through alterations in upstream regulatory pathways. Inflammatory signaling pathways, such as MAPK and PI3K/AKT, can interact with NF-κB and influence HHEX activity [35]. Cross-talk between these pathways can modulate HHEX function by altering its phosphorylation status, subcellular localization, or interaction with co-factors required for transcriptional regulation. Chronic inflammation involves the infiltration of immune cells, such as macrophages and lymphocytes, into thyroid tissue. These immune cells produce cytokines and other mediators that affect HHEX expression and function, contributing to thyroid dysfunction and disease progression. The potential mechanisms of HHEX dysregulation in thyroid inflammation include direct inhibition by NF-κB, which inhibits HHEX transcription by directly binding to regulatory regions or by recruiting repressive chromatin-modifying complexes; epigenetic modifications, where inflammatory cytokines and oxidative stress induce changes in the HHEX promoter region, altering its accessibility to transcriptional machinery; post-translational modifications, where oxidative stress and inflammatory signaling pathways lead to HHEX protein modifications that affect its stability and functional activity; and disruption of signaling networks, where inflammation disrupts signaling networks that regulate HHEX expression and function, impairing its ability to coordinate thyroid morphogenesis and hormone synthesis. Inflammation, particularly through NF-κB activation, can dysregulate HHEX in multiple ways, leading to the disruption of thyroid development and function. This dysregulation contributes to thyroid disorders, including thyroiditis and potentially thyroid cancer [36].

##### Resultant dysregulation in thyroid cell fate and resultant impact on FTC development

NF-κB-mediated disruption of HHEX (Hematopoietically-expressed homeobox protein) can significantly impact thyroid follicular cell fate and predispose the thyroid gland to follicular thyroid carcinoma (FTC). Chronic inflammation in the thyroid gland, such as autoimmune thyroiditis or chronic infection, leads to sustained activation of NF-κB. Pro-inflammatory cytokines, such as TNF-α and IL-1β, stimulate the NF-κB pathway by inducing the degradation of IκB proteins, allowing NF-κB dimers to translocate into the nucleus and modulate gene expression. NF-κB can directly inhibit HHEX expression by binding to NF-κB response elements in the HHEX promoter region, thereby reducing its transcriptional activity [37]. NF-κB activation may also induce the expression of microRNAs or other factors that target HHEX mRNA stability, leading to decreased HHEX protein levels. Additionally, NF-κB signaling can induce changes in chromatin structure, such as histone modifications and DNA methylation, that silence HHEX expression, further impairing its function. HHEX is essential for thyroid follicular cell differentiation and function by regulating the expression of genes involved in thyroid hormone synthesis, such as thyroglobulin and thyroperoxidase. Proper HHEX activity ensures the maintenance of differentiated thyroid follicular cell identity and function. NF-κB-mediated inhibition of HHEX results in reduced expression of thyroid-specific genes, leading to a loss of differentiated cell markers [38]. Decreased HHEX levels can promote dedifferentiation of thyroid follicular cells, making them more prone to acquiring oncogenic mutations and phenotypes associated with cancer. Dysregulation of HHEX due to NF-κB activation can initiate the molecular events leading to FTC development. NF-κB signaling supports uncontrolled cell proliferation by upregulating genes involved in cell cycle progression and inhibiting apoptosis. Chronic inflammation and oxidative stress associated with NF-κB activation can induce DNA damage and genomic instability, promoting oncogenic transformations in thyroid cells. The mechanisms of FTC development include the loss of differentiated thyroid follicular cell characteristics, a hallmark of thyroid cancer progression. NF-κB activation enhances cell survival mechanisms by upregulating anti-apoptotic proteins and promoting resistance to cell death signals. NF-κB-mediated inflammation creates a pro-tumorigenic microenvironment characterized by increased angiogenesis, immune evasion, and altered metabolic pathways. NF-κB-mediated disruption of HHEX in thyroid follicular cells disrupts cell fate determination, leading to dedifferentiation and a predisposition to follicular thyroid carcinoma (FTC). This dysregulation alters thyroid-specific gene expression, impairs hormone synthesis, and promotes cellular processes associated with cancer development [39, 40].

#### 5. TSH (Thyroid-Stimulating Hormone) signaling pathway Role of NF-κB in disrupting it

The TSH (Thyroid-Stimulating Hormone) signaling pathway is essential for thyroid development, function, and homeostasis. Dysregulation of this pathway due to inflammation, particularly involving NF-κB activation, can profoundly impact thyroid health and contribute to thyroid disorders [41]. TSH, produced by the pituitary gland, binds to its receptor (TSHR) on thyroid follicular cells, stimulating thyroid hormone synthesis and secretion. Key components include TSHR activation, which leads to increased intracellular cAMP levels and subsequently activates protein kinase A (PKA). PKA then phosphorylates transcription factors like CREB, regulating gene expression involved in thyroid function. Inflammatory conditions elevate cytokines such as TNF-α, IL-1β, and IL-6 in the thyroid gland, interfering with TSH signaling by disrupting TSHR expression, altering cAMP production, or modifying downstream signaling pathways. NF-κB, a key mediator of inflammation activated by pro-inflammatory cytokines and cellular stress, can inhibit TSHR gene transcription directly or indirectly through downstream effectors, reducing the responsiveness of thyroid cells to TSH stimulation [42]. NF-κB signaling can also interfere with adenylate cyclase activity or cAMP synthesis, impairing the generation of intracellular signals necessary for thyroid hormone production. Additionally, NF-κB activation alters the expression of genes involved in thyroid differentiation and function, potentially disrupting the TSH-mediated transcriptional program. Inflammation induces oxidative stress in thyroid cells through the production of reactive oxygen species (ROS), which can affect components of the TSH signaling pathway, including TSHR and adenylate cyclase, leading to impaired cAMP signaling and thyroid hormone synthesis. Immune cells infiltrate the thyroid during chronic inflammation, secreting cytokines and chemokines that directly impact TSH signaling components, contributing to thyroid dysfunction and disease progression [43].

Dysregulated TSH signaling results in reduced thyroid hormone production, leading to hypothyroidism or subclinical hypothyroidism. The disruption of TSH signaling can also impair the differentiation of thyroid follicular cells, contributing to thyroid nodules or goiter formation. Persistent inflammation and dysregulated TSH signaling create a pro-tumorigenic microenvironment in the thyroid gland, promoting cell proliferation and inhibiting apoptosis. This contributes to the development of thyroid cancers such as papillary thyroid carcinoma. Inflammation, particularly through NF-κB activation, can disrupt the TSH signaling pathway in multiple ways, impacting thyroid hormone synthesis, cell differentiation, and predisposing the thyroid gland to disorders such as hypothyroidism and thyroid cancer [44].

##### Resultant dysregulation in thyroid cell fate and resultant impact on FTC development

NF-κB-mediated disruption of the TSH (Thyroid-Stimulating Hormone) signaling pathway can have significant implications for thyroid follicular cell fate and predispose the thyroid gland to follicular thyroid carcinoma (FTC). TSH normally binds to its receptor (TSHR) on thyroid follicular cells, initiating a signaling cascade that includes the activation of adenylate cyclase, increased intracellular cyclic AMP (cAMP) levels, and subsequent phosphorylation of transcription factors such as CREB. This pathway is essential for regulating thyroid hormone synthesis and maintaining thyroid function [45]. However, NF-κB activation can interfere with this signaling process in several ways. It can inhibit TSHR gene transcription directly by binding to NF-κB response elements in the TSHR promoter region, leading to reduced expression of TSHR on thyroid follicular cells. This decreases their responsiveness to TSH stimulation, resulting in reduced iodine uptake, impaired thyroid hormone synthesis, and disrupted thyroid follicular cell differentiation [46]. Additionally, NF-κB signaling can interfere with adenylate cyclase activity or disrupt cAMP synthesis pathways in thyroid cells. Lower cAMP levels result in decreased activation of protein kinase A (PKA) and CREB, which impairs the transcriptional machinery necessary for thyroid hormone synthesis and cell differentiation. Furthermore, NF-κB activation alters the expression of genes involved in thyroid differentiation and function, potentially suppressing the expression of thyroid-specific genes under TSH control. This disrupted transcriptional regulation leads to a loss of thyroid follicular cell identity and differentiation markers, promoting a more proliferative and less differentiated phenotype. The consequences of NF-κB-mediated disruption of TSH signaling are profound. The inhibition of TSH signaling disrupts the differentiation program of thyroid follicular cells, leading to a dedifferentiated state. Dedifferentiated thyroid cells exhibit increased proliferative capacity due to dysregulated signaling pathways and reduced responsiveness to growth inhibitory signals. Chronic inflammation and oxidative stress associated with NF-κB activation can also induce DNA damage and genomic instability, contributing to the acquisition of oncogenic mutations [47]. The dysregulation of TSH signaling by NF-κB creates a favorable microenvironment for the initiation of thyroid cancer. NF-κB activation enhances anti-apoptotic pathways and survival signals, protecting thyroid cells from programmed cell death. Additionally, inflammatory cytokines induced by NF-κB promote angiogenesis and facilitate the metastatic spread of thyroid cancer cells [48].

#### 6. RET/PTC

##### Role of NF-κB in disrupting it

RET/PTC (Rearranged during Transfection/Papillary Thyroid Carcinoma) gene rearrangements are oncogenic drivers frequently observed in papillary thyroid carcinoma (PTC). These genetic alterations result from chromosomal rearrangements that fuse the RET proto-oncogene with various partner genes, leading to constitutive activation of RET tyrosine kinase activity [49]. Inflammation, especially involving NF-κB activation, can significantly influence RET/PTC oncogene expression and function through several mechanisms. NF-κB activation, in response to chronic inflammation, can induce genetic instability by producing reactive oxygen species (ROS) and causing DNA damage, which promotes chromosomal rearrangements involving RET and its fusion partners. This activation also leads to the upregulation of pro-inflammatory cytokines such as TNF-α and IL-6, which can promote cell proliferation and survival, thereby contributing to the clonal expansion of thyroid cells harboring RET/PTC rearrangements [50]. Additionally, NF-κB enhances cell survival pathways by promoting the expression of anti-apoptotic proteins like Bcl-2 and Bcl-xL, as well as inhibitors of apoptosis proteins (IAPs), which protect RET/PTC-transformed cells from apoptotic death and support their survival and proliferation. The consequences of NF-κB activation on RET/PTC are significant. NF-κB can directly enhance the transcriptional activity of RET/PTC fusion genes by binding to NF-κB response elements within their regulatory regions [51]. This results in sustained activation of RET tyrosine kinase signaling pathways, which promotes cell growth and survival. Furthermore, NF-κB-mediated activation of signaling pathways such as MAPK and PI3K/AKT synergizes with RET/PTC oncogenic signaling, amplifying the oncogenic potential of thyroid follicular cells and driving tumor progression. NF-κB also alters the expression of genes involved in cell cycle regulation, angiogenesis, and metastasis, which contributes to the aggressive phenotype of RET/PTC-driven thyroid cancers [52].

##### Resultant dysregulation in thyroid cell fate and resultant impact on FTC development

NF-κB activation significantly impacts RET/PTC (Rearranged during Transfection/Papillary Thyroid Carcinoma) gene rearrangements, playing a key role in disrupting thyroid follicular cell fate and increasing the risk of follicular thyroid carcinoma (FTC). NF-κB activation is triggered in response to pro-inflammatory cytokines such as TNF-α and IL-6, which are elevated during thyroid inflammation [53]. This process results in the translocation of NF-κB to the nucleus, where it regulates the expression of genes involved in inflammation, cell survival, and proliferation. Chronic inflammation in the thyroid gland also leads to the generation of reactive oxygen species (ROS) and oxidative stress. NF-κB activation, in response to oxidative stress, amplifies inflammatory signaling pathways, contributing to cellular damage and transformation. Furthermore, inflammatory conditions lead to the recruitment of immune cells, such as macrophages and lymphocytes, to the thyroid gland. These immune cells secrete cytokines and growth factors that activate NF-κB, creating a pro-inflammatory microenvironment conducive to tumor development. The impact of NF-κB on RET/PTC and thyroid follicular cell fate is profound [54]. NF-κB activation can directly enhance the transcriptional activity of RET/PTC fusion genes by binding to NF-κB response elements within their regulatory regions. This results in sustained activation of RET tyrosine kinase signaling pathways, which promotes cell growth and survival. RET/PTC oncogenes disrupt normal thyroid follicular cell differentiation programs, and NF-κB-mediated upregulation of RET/PTC exacerbates this disruption, leading to the loss of differentiated cell characteristics and promoting a more dedifferentiated and aggressive phenotype. Additionally, NF-κB activation promotes cell proliferation and inhibits apoptosis through the activation of downstream signaling pathways such as MAPK and PI3K/AKT. These pathways synergize with RET/PTC oncogenic signaling, enhancing cellular proliferation, survival, and resistance to apoptosis. NF-κB-mediated activation of RET/PTC contributes to oncogenic transformation of thyroid follicular cells, which is essential for the initiation and progression of follicular thyroid carcinoma [55]. Chronic NF-κB activation also induces DNA damage and genomic instability, creating a mutagenic environment that facilitates the accumulation of additional genetic alterations necessary for tumor progression. Moreover, NF-κB activation promotes an inflammatory and immunosuppressive tumor microenvironment that supports tumor growth, angiogenesis, and metastasis [56].

#### 7. MAPK/ERK pathway

##### Role of NF-κB in disrupting it

MAPK/ERK pathway, key in thyroid development, regulates various processes such as cell proliferation, differentiation, and survival. This pathway is activated when growth factors, cytokines, and TSH bind to receptor tyrosine kinases (RTKs) on thyroid follicular cells. Activation of RTKs recruits and activates Ras, which in turn activates Raf kinase. Raf then phosphorylates and activates MEK (MAPK/ERK kinase), which subsequently phosphorylates and activates ERK1/2 (Extracellular Signal-Regulated Kinase) [57]. Activated ERK1/2 translocates to the nucleus, where it phosphorylates transcription factors like Elk-1 and CREB, regulating gene expression related to cell proliferation, differentiation, and survival. Inflammation, particularly through NF-κB activation, can disrupt the MAPK/ERK pathway significantly. NF-κB is activated by pro-inflammatory cytokines such as TNF-α, IL-1β, and IL-6, which are elevated in thyroid inflammation. These cytokines facilitate NF-κB translocation to the nucleus, where it regulates genes involved in inflammation and cellular responses. Inflammatory conditions also induce oxidative stress in thyroid cells, leading to the production of reactive oxygen species (ROS). ROS can activate Ras and Raf kinases directly or through oxidative modifications, thereby enhancing MAPK/ERK pathway signaling. NF-κB activation and cytokine signaling can lead to sustained activation of RTKs and Ras, resulting in prolonged activation of the MAPK/ERK pathway. This persistent activation can alter gene expression profiles, promoting cell proliferation and survival, and contributing to thyroid hyperplasia or neoplasia [58]. Additionally, NF-κB can modulate the expression of genes encoding components of the MAPK/ERK pathway, such as RTKs, Ras, and Raf. Changes in expression levels or mutations in these genes can dysregulate pathway activity, leading to thyroid dysfunction and disease progression. Inflammatory mediators can also disrupt negative feedback mechanisms that normally regulate MAPK/ERK pathway activity, causing sustained ERK1/2 activation and promoting aberrant cell growth and survival in thyroid cells [59]. The consequences of such dysregulation include thyroid hyperplasia, where persistent activation of the MAPK/ERK pathway leads to thyroid follicular cell hyperplasia and goiter formation. Additionally, dysregulated MAPK/ERK pathway signaling is associated with thyroid cancer development, including papillary thyroid carcinoma (PTC). Altered MAPK/ERK signaling can also disrupt thyroid hormone synthesis and secretion, contributing to conditions such as hypothyroidism or thyroid hormone resistance [60].

##### Resultant dysregulation in thyroid cell fate and resultant impact on FTC development

NF-κB activation can significantly disrupt thyroid follicular cell fate and predispose the thyroid gland to follicular thyroid carcinoma (FTC) by influencing the MAPK/ERK (Mitogen-Activated Protein Kinase/Extracellular Signal-Regulated Kinase) pathway. Pro-inflammatory cytokines such as TNF-α and IL-1β stimulate NF-κB activation in response to thyroid inflammation. This activation causes NF-κB to translocate to the nucleus, where it regulates gene expression involved in inflammation, cell survival, and proliferation. NF-κB activation also enhances the expression and activity of receptor tyrosine kinases (RTKs) and downstream components of the MAPK/ERK pathway. Increased RTK activation results in sustained phosphorylation and activation of the Raf-MEK-ERK cascades, promoting cell proliferation and survival [61]. Furthermore, NF-κB disrupts negative feedback loops that normally regulate MAPK/ERK pathway activity, leading to persistent ERK1/2 activation. This disruption alters gene expression profiles and promotes thyroid follicular cell proliferation and survival. Prolonged activation of the MAPK/ERK pathway by NF-κB can disrupt thyroid follicular cell differentiation programs, resulting in the loss of normal thyroid follicular cell characteristics and differentiation markers [62]. This promotes a dedifferentiated state typical of thyroid cancers.

Additionally, NF-κB-mediated activation of the MAPK/ERK pathway enhances cell proliferation and inhibits apoptosis in thyroid follicular cells, leading to increased cell proliferation combined with impaired differentiation. This combination predisposes thyroid cells to oncogenic transformation and the development of follicular thyroid carcinoma. Dysregulated MAPK/ERK signaling, driven by NF-κB activation, promotes the accumulation of genetic alterations and genomic instability in thyroid follicular cells, creating a favorable environment for the initiation and progression of follicular thyroid carcinoma. Activated MAPK/ERK signaling enhances cell cycle progression, angiogenesis, and resistance to apoptosis, facilitating the growth and metastasis of thyroid cancer cells. As a result, FTC arises from thyroid follicular cells that have undergone oncogenic transformation due to sustained NF-κB-mediated MAPK/ERK pathway activation [63, 64].

#### 8. PI3K/AKT pathway

##### Role of NF-κB in disrupting it

The PI3K/AKT pathway (Phosphoinositide 3-kinase/Protein Kinase B pathway) plays a key role in thyroid development by regulating cell growth, survival, metabolism, and differentiation. This pathway is typically activated by growth factors such as insulin-like growth factor 1 (IGF-1) and insulin, which act on receptor tyrosine kinases (RTKs) or G protein-coupled receptors (GPCRs) on thyroid follicular cells [65]. Activation of RTKs or GPCRs leads to the recruitment and activation of PI3K, which then phosphorylates phosphatidylinositol 4,5-bisphosphate (PIP2) to produce phosphatidylinositol 3,4,5-trisphosphate (PIP3). This process recruits AKT (also known as Protein Kinase B, PKB) to the cell membrane, where AKT is phosphorylated and activated. Activated AKT then phosphorylates downstream targets involved in cell survival, such as Bad and FOXO, as well as targets related to proliferation, such as mTOR, and metabolism, such as GSK3. Inflammation can disrupt this pathway, particularly through the activation of NF-κB. Pro-inflammatory cytokines like TNF-α and IL-1β stimulate NF-κB activation in response to thyroid inflammation [66]. NF-κB translocates to the nucleus and induces the expression of genes involved in inflammation and cellular responses. Additionally, inflammatory conditions lead to oxidative stress in thyroid cells, resulting in the production of reactive oxygen species (ROS). ROS can activate RTKs directly or through oxidative modifications, thereby enhancing PI3K/AKT pathway signaling. The impact of NF-κB activation on the PI3K/AKT pathway includes enhanced signaling through RTKs or GPCRs, leading to increased pathway activation. Sustained AKT activation promotes cell survival, proliferation, and metabolism, which can contribute to thyroid cell hyperplasia or neoplasia under inflammatory conditions. NF-κB can also modulate the expression of genes encoding components of the PI3K/AKT pathway, such as RTKs, PI3K subunits, and AKT isoforms. Altered expression levels or mutations in these genes can disrupt pathway activity, promoting thyroid dysfunction and disease progression. Furthermore, inflammatory mediators can disrupt negative feedback mechanisms that normally regulate the PI3K/AKT pathway, resulting in sustained AKT activation and altered gene expression profiles that promote thyroid follicular cell growth and survival [67]. The consequences of dysregulated PI3K/AKT signaling include thyroid hyperplasia, which can contribute to goiter formation, and thyroid cancer development, including follicular and papillary thyroid carcinomas. Altered PI3K/AKT signaling can also disrupt thyroid hormone synthesis and secretion, leading to hypothyroidism or thyroid hormone resistance [68].

##### Resultant dysregulation in thyroid cell fate and resultant impact on FTC development

NF-κB activation and its impact on the PI3K/AKT pathway can disrupt thyroid follicular cell fate and predispose the thyroid gland to follicular thyroid carcinoma (FTC). Pro-inflammatory cytokines such as TNF-α and IL-1β stimulate NF-κB activation in response to thyroid inflammation. NF-κB then translocates to the nucleus, inducing the expression of genes involved in inflammation, cell survival, and proliferation. This activation enhances the expression and activity of receptor tyrosine kinases (RTKs) and G protein-coupled receptors (GPCRs) on thyroid follicular cells, leading to increased PI3K activity. PI3K phosphorylates phosphatidylinositol 4,5-bisphosphate (PIP2) to form phosphatidylinositol 3,4,5-trisphosphate (PIP3), which recruits AKT (Protein Kinase B) to the cell membrane. Activated AKT then promotes cell survival, proliferation, and metabolism by phosphorylating downstream targets such as mTOR (mammalian Target of Rapamycin) and FOXO (Forkhead box O) transcription factors [69]. The persistent activation of the PI3K/AKT pathway due to NF-κB influence can disrupt thyroid follicular cell differentiation programs, leading to the loss of normal thyroid follicular cell characteristics and a dedifferentiated state conducive to thyroid cancer development. Enhanced PI3K/AKT signaling supports thyroid cell growth and survival, increasing cell proliferation while inhibiting apoptosis. This combination of increased proliferation and impaired differentiation predisposes thyroid cells to oncogenic transformation, favoring the development of follicular thyroid carcinoma. Dysregulated PI3K/AKT signaling driven by NF-κB activation promotes genomic instability and the accumulation of genetic alterations in thyroid follicular cells [70]. This creates a favorable environment for the initiation and progression of follicular thyroid carcinoma. Activated PI3K/AKT signaling also enhances cell cycle progression, angiogenesis, and resistance to apoptosis, facilitating the growth and metastasis of thyroid cancer cells. Consequently, FTC arises from thyroid follicular cells that have undergone oncogenic transformation due to sustained NF-κB-mediated PI3K/AKT pathway activation [71, 72].

#### 9. Wnt/β-catenin signaling pathway Role of NF-κB in disrupting it

The Wnt/β-catenin signaling pathway plays a key role in thyroid development, regulating processes such as cell proliferation, differentiation, and tissue morphogenesis. When functioning normally, Wnt ligands bind to Frizzled receptors and co-receptors like LRP5/6, stabilizing β-catenin in the cytoplasm [73]. Stabilized β-catenin then translocates into the nucleus, where it interacts with TCF/LEF transcription factors to regulate the expression of genes involved in cell proliferation, survival, and differentiation, such as c-Myc, Cyclin D1, and Axin2. However, dysregulation of the Wnt/β-catenin pathway due to inflammation, including NF-κB activation, can significantly impact thyroid function and contribute to thyroid disorders. Inflammatory cytokines like TNF-α and IL-1β stimulate NF-κB activation in response to thyroid inflammation [74]. NF-κB translocates to the nucleus, inducing the expression of genes involved in inflammation and cellular responses. Inflammatory conditions also induce oxidative stress in thyroid cells, leading to the production of reactive oxygen species (ROS). These ROS can activate Wnt signaling through mechanisms such as stabilization of β-catenin and modulation of Wnt ligand expression. The activation of NF-κB can enhance Wnt signaling by either increasing Wnt ligand expression or directly stabilizing β-catenin, which promotes sustained Wnt/β-catenin pathway activation. This can lead to excessive cell proliferation, survival, and altered differentiation in thyroid cells. NF-κB can also modulate the expression of genes involved in regulating the Wnt/β-catenin pathway, such as Wnt ligands, receptors, and intracellular components. Altered expression levels or mutations in these genes can disrupt pathway activity, contributing to thyroid dysfunction and disease progression. Furthermore, inflammatory mediators can interfere with negative feedback mechanisms that normally regulate Wnt/β-catenin pathway activity, leading to sustained β-catenin accumulation in the nucleus, which alters gene expression profiles and promotes abnormal cell behaviors in thyroid follicular cells [75]. The consequences of such dysregulation include thyroid hyperplasia, where persistent activation of the Wnt/β-catenin pathway leads to the hyperplasia of thyroid follicular cells and contributes to goiter formation. Additionally, dysregulated Wnt/β-catenin signaling is associated with the development of thyroid cancer, including follicular thyroid carcinoma (FTC) and thyroid adenomas. Altered signaling can also disrupt thyroid hormone synthesis and secretion, leading to hypothyroidism or thyroid hormone resistance [76].

##### Resultant dysregulation in thyroid cell fate and resultant impact on FTC development

NF-κB activation and its impact on the Wnt/β-catenin signaling pathway can disrupt thyroid follicular cell fate and increase the risk of follicular thyroid carcinoma (FTC). Inflammatory cytokines such as TNF-α and IL-1β trigger NF-κB activation in response to thyroid inflammation. This activation leads NF-κB to translocate to the nucleus, where it induces the expression of genes involved in inflammation, cell survival, and proliferation. Additionally, NF-κB activation can enhance the expression of Wnt ligands like Wnt1 and Wnt3a in thyroid follicular cells. Elevated levels of these Wnt ligands activate the Wnt/β-catenin pathway by stabilizing β-catenin in the cytoplasm and facilitating its translocation to the nucleus [77]. NF-κB-induced oxidative stress or inflammatory mediators can further stabilize β-catenin by inhibiting its degradation complex, which includes components such as APC and GSK-3β. This stabilization results in the accumulation of β-catenin in the nucleus, where it interacts with TCF/LEF transcription factors to activate Wnt target genes involved in cell proliferation and survival. The persistent activation of the Wnt/β-catenin pathway under NF-κB influence can disrupt normal thyroid follicular cell differentiation programs, leading to a dedifferentiated state characterized by the loss of typical follicular cell markers and enhanced stem-like properties, which are conducive to oncogenic transformation [78]. Furthermore, the activation of the Wnt/β-catenin pathway promotes increased thyroid cell proliferation and inhibits apoptosis. This combination of heightened proliferation and impaired differentiation predisposes thyroid cells to oncogenic transformation, thereby favoring the development of follicular thyroid carcinoma. Dysregulated Wnt/β-catenin signaling, driven by NF-κB activation, leads to genomic instability and the accumulation of genetic alterations in thyroid follicular cells, creating a permissive environment for FTC initiation and progression [79]. The activated Wnt/β-catenin signaling also enhances cell cycle progression, angiogenesis, and resistance to apoptosis, facilitating the growth and metastasis of thyroid cancer cells. Consequently, FTC arises from thyroid follicular cells that have undergone oncogenic transformation due to sustained NF-κB-mediated Wnt/β-catenin pathway activation [80].

#### 10. FGF

##### Role of NF-κB in disrupting it

Fibroblast Growth Factors (FGFs) play essential roles in thyroid development, influencing cell proliferation, differentiation, and tissue morphogenesis. FGFs, through their receptors (FGFRs), activate downstream signaling pathways such as MAPK/ERK and PI3K/AKT, promoting thyroid follicular cell proliferation and survival. Additionally, FGF signaling is essential for regulating the differentiation of thyroid progenitor cells into functional thyroid follicular cells, which produce thyroid hormones [81]. Dysregulation of FGF signaling due to inflammation, including NF-κB activation, can significantly impact thyroid function and contribute to thyroid disorders. Inflammatory cytokines, such as TNF-α and IL-1β, stimulate NF-κB activation in response to thyroid inflammation. This activation leads NF-κB to translocate to the nucleus and induce the expression of genes involved in inflammation and cellular responses, including those that regulate FGF signaling [82]. Furthermore, inflammatory conditions induce oxidative stress in thyroid cells, which can modulate receptor tyrosine kinase activities, including FGFRs. NF-κB activation can regulate the expression of FGFRs and their isoforms on thyroid follicular cells. Altered FGFR expression levels can affect the sensitivity of thyroid cells to FGF ligands, influencing cell proliferation and differentiation. Inflammatory cytokines and oxidative stress can also influence the expression of FGF ligands within the thyroid microenvironment. Changes in FGF ligand expression patterns can disrupt the balance of signaling cues required for proper thyroid development and function. NF-κB-mediated inflammation can cross-talk with FGF signaling pathways, such as MAPK/ERK and PI3K/AKT, leading to dysregulated FGF signaling pathways [83]. This dysregulation contributes to aberrant thyroid cell behaviors, including proliferation, survival, and differentiation, which are characteristic of thyroid disorders and cancer. The consequences of dysregulated FGF signaling under inflammatory conditions include thyroid follicular cell hyperplasia, which can contribute to goiter formation. Aberrant FGF signaling is also associated with thyroid cancer development, including follicular thyroid carcinoma (FTC) and papillary thyroid carcinoma (PTC). Moreover, altered FGF signaling disrupts thyroid hormone synthesis and secretion, contributing to hypothyroidism or thyroid hormone resistance.

Additionally, assessing FGF signaling pathway activation in thyroid tumors can serve as a prognostic indicator for disease aggressiveness and response to targeted therapies. Overall, NF-κB activation disrupts FGF signaling in thyroid follicular cells, affecting essential processes for normal thyroid function. Dysregulated FGF signaling under inflammatory conditions contributes to thyroid disorders and cancer by promoting abnormal cell behaviors and oncogenic transformation. Further research into the interplay between NF-κB and FGF signaling pathways is critical for developing effective therapeutic strategies and improving clinical outcomes in patients with thyroid diseases [84].

##### Resultant dysregulation in thyroid cell fate and resultant impact on FTC development

NF-κB activation can significantly impact the FGF (Fibroblast Growth Factor) signaling pathway in thyroid follicular cells, potentially disrupting cell fate regulation and increasing the risk of follicular thyroid carcinoma (FTC). Inflammatory cytokines, such as TNF-α and IL-1β, induce NF-κB activation in response to thyroid inflammation. This activation causes NF-κB to translocate to the nucleus and regulate the expression of genes involved in inflammation and cellular responses, including those related to FGF signaling [85]. NF-κB activation can influence the expression levels of FGFRs (Fibroblast Growth Factor Receptors) on thyroid follicular cells. Altered FGFR expression affects the responsiveness of thyroid cells to FGF ligands, impacting cell proliferation, survival, and differentiation. Additionally, NF-κB-mediated inflammation can interact with intracellular signaling pathways activated by FGFs, such as MAPK/ERK and PI3K/AKT. Dysregulated FGF signaling pathways can disrupt normal thyroid follicular cell fate regulation, leading to abnormal proliferation and impaired differentiation [86]. Sustained activation of NF-κB and dysregulated FGF signaling can disrupt the normal differentiation programs of thyroid follicular cells, leading to a dedifferentiated state where thyroid cells lose their specialized functions and acquire stem-like properties, which contributes to tumorigenesis. Furthermore, NF-κB-mediated dysregulation of FGF signaling pathways promotes thyroid cell proliferation and inhibits apoptosis. Enhanced cell proliferation combined with impaired differentiation creates a cellular environment prone to oncogenic transformation, particularly towards follicular thyroid carcinoma.

Dysregulated FGF signaling, influenced by NF-κB activation, contributes to genomic instability and accumulation of genetic alterations in thyroid follicular cells, creating a favorable environment for the initiation and progression of follicular thyroid carcinoma. Aberrant FGF signaling under the influence of NF-κB supports tumor growth by enhancing cell proliferation, angiogenesis, and resistance to apoptosis. FTC arises from thyroid follicular cells that have undergone oncogenic transformation due to sustained NF-κB-mediated disruption of FGF signaling pathways [87, 88].

#### 11. Sonic Hedgehog (Shh) signaling Role of NF-κB in disrupting it

Sonic Hedgehog (Shh) signaling is significant in embryonic development, including thyroid development, where it influences cell differentiation, proliferation, and tissue patterning. Shh signaling plays a role in specifying thyroid progenitor cells and promoting their differentiation into functional thyroid follicular cells. It also regulates cell proliferation and tissue morphogenesis in the developing thyroid gland, ensuring proper growth and structure. Dysregulation of Shh signaling due to inflammation, including NF-κB activation, can impact thyroid function and contribute to thyroid disorders. Inflammatory cytokines such as TNF-α and IL-1β induce NF-κB activation in response to thyroid inflammation [89]. This activation causes NF-κB to translocate to the nucleus, where it regulates the expression of genes involved in inflammation and cellular responses, potentially affecting Shh signaling components. Inflammatory conditions can also induce oxidative stress in thyroid cells, modulating signaling pathways involved in cell fate determination, including Shh. NF-κB activation can influence the expression of Shh ligands and receptors on thyroid follicular cells, leading to altered Shh expression levels [90]. This change can affect the sensitivity of thyroid cells to Shh ligands and perturb downstream signaling pathways. Moreover, NF-κB-mediated inflammation can interact with intracellular signaling pathways activated by Shh, such as the Smoothened (Smo)-Glioma-associated oncogene homolog (Gli) pathway. Dysregulated Shh signaling disrupts normal thyroid follicular cell fate regulation, potentially leading to abnormal proliferation, differentiation, or both. Dysregulated Shh signaling under inflammatory conditions can result in abnormal thyroid gland development, manifesting as either reduced or excessive tissue growth (hypoplasia or hyperplasia) [91]. Aberrant Shh signaling is also associated with thyroid cancer development, including medullary thyroid carcinoma (MTC) and possibly other types through complex interactions with other pathways. Altered Shh signaling disrupts thyroid hormone synthesis and secretion, contributing to hypothyroidism or thyroid hormone resistance [92].

##### Resultant dysregulation in thyroid cell fate and resultant impact on FTC development

NF-κB activation can significantly impact Sonic Hedgehog (Shh) signaling in thyroid follicular cells, potentially disrupting cell fate regulation and contributing to the predisposition of the thyroid gland to follicular thyroid carcinoma (FTC). Inflammatory cytokines, such as TNF-α and IL-1β, induce NF-κB activation in response to thyroid inflammation [93]. This activation causes NF-κB to translocate to the nucleus, where it regulates the expression of genes involved in inflammation and cellular responses, which can affect components of the Shh signaling pathway. NF-κB activation can also influence the expression levels of Shh ligands and receptors on thyroid follicular cells.

Altered Shh expression levels can disrupt the normal signaling cascade, affecting downstream events involved in cell fate determination, proliferation, and differentiation. Furthermore, NF-κB-mediated inflammation can cross-talk with intracellular signaling pathways activated by Shh, such as the Smoothened (Smo)-Glioma-associated oncogene homolog (Gli) pathway [94]. Dysregulated Shh signaling can lead to aberrant activation of Gli transcription factors, influencing the expression of target genes involved in cell cycle regulation and cell fate decisions. Dysregulated Shh signaling under NF-κB influence can disrupt the normal differentiation programs of thyroid follicular cells. This disruption may lead to a dedifferentiated state where thyroid cells lose their specialized functions and acquire stem-like properties, which are conducive to oncogenic transformation. Additionally, activation of Shh signaling by NF-κB can enhance thyroid cell proliferation and survival mechanisms. Enhanced cell proliferation combined with impaired differentiation creates a cellular environment prone to oncogenic transformation, particularly towards follicular thyroid carcinoma. Dysregulated Shh signaling, influenced by NF-κB activation, promotes genomic instability and accumulation of genetic alterations in thyroid follicular cells. These molecular changes create a favorable milieu for the initiation and progression of follicular thyroid carcinoma [95]. Aberrant Shh signaling under NF-κB influence supports tumor growth by enhancing cell proliferation, angiogenesis, and resistance to apoptosis. FTC arises from thyroid follicular cells that have undergone oncogenic transformation due to sustained NF-κB-mediated disruption of Shh signaling pathways [96].

#### 12. Bone Morphogenetic Proteins (BMPs) Role of NF-κB in disrupting it

Bone Morphogenetic Proteins (BMPs) play critical roles in thyroid development by influencing processes such as cell differentiation, proliferation, and tissue patterning. BMP signaling is essential for regulating the differentiation of thyroid progenitor cells into functional thyroid follicular cells and for maintaining proper tissue morphogenesis and growth during thyroid gland development [97]. Dysregulation of BMP signaling due to inflammation, including NF-κB activation, can impact thyroid function and contribute to thyroid disorders. Inflammatory cytokines, such as TNF-α and IL-1β, induce NF-κB activation in response to thyroid inflammation. This activation causes NF-κB to translocate to the nucleus, where it regulates the expression of genes involved in inflammation and cellular responses, which can potentially affect BMP signaling components. Inflammatory conditions also induce oxidative stress in thyroid cells, which can modulate BMP signaling pathways involved in cell fate determination and proliferation. NF-κB activation can influence the expression levels of BMP ligands, such as BMP2 and BMP4, and their receptors, such as BMPR1A and BMPR2, on thyroid follicular cells. Altered BMP expression levels can disrupt the balance of BMP signaling, affecting downstream signaling events essential for thyroid cell differentiation and function. Additionally, NF-κB-mediated inflammation can cross-talk with intracellular signaling pathways activated by BMPs, such as SMAD-dependent and SMAD-independent pathways [98]. This dysregulation can lead to aberrant activation of downstream effectors, influencing gene expression profiles and cellular responses in thyroid follicular cells. Dysregulated BMP signaling under inflammatory conditions can result in abnormal thyroid gland development, manifesting as either reduced or excessive tissue growth, known as hypoplasia or hyperplasia. Aberrant BMP signaling is also associated with thyroid cancer development, including papillary thyroid carcinoma (PTC) and potentially other types through interactions with other pathways. Moreover, altered BMP signaling disrupts thyroid hormone synthesis and secretion, contributing to hypothyroidism or thyroid hormone resistance [99, 100].

##### Resultant dysregulation in thyroid cell fate and resultant impact on FTC development

NF-κB activation can impact Bone Morphogenetic Protein (BMP) signaling in thyroid follicular cells, potentially disrupting cell fate regulation and increasing the risk of developing follicular thyroid carcinoma (FTC). Inflammatory cytokines such as TNF-α and IL-1β trigger NF-κB activation in response to thyroid inflammation. This process leads NF-κB to translocate to the nucleus, where it regulates the expression of genes involved in inflammation and cellular responses, which may in turn affect BMP signaling components. NF-κB activation can influence the expression levels of BMP ligands, like BMP2 and BMP4, and their receptors, such as BMPR1A and BMPR2, on thyroid follicular cells.

Changes in BMP expression levels can disrupt the balance of BMP signaling, affecting downstream signaling events that are essential for thyroid cell differentiation and function [101]. Moreover, NF-κB-mediated inflammation can interact with intracellular signaling pathways activated by BMPs, including both SMAD-dependent and SMAD-independent pathways. This dysregulation can result in aberrant activation of downstream effectors, altering gene expression profiles and cellular responses in thyroid follicular cells. Under the influence of NF-κB, dysregulated BMP signaling can lead to a loss of normal differentiation programs in thyroid follicular cells. This disruption may cause cells to enter a dedifferentiated state where they lose their specialized functions and gain stem-like properties, which can promote oncogenic transformation. Additionally, NF-κB-induced activation of BMP signaling can enhance thyroid cell proliferation and survival mechanisms [102]. The combination of increased cell proliferation and impaired differentiation fosters an environment conducive to oncogenic transformation, particularly towards follicular thyroid carcinoma. Dysregulated BMP signaling, influenced by NF-κB activation, can lead to genomic instability and the accumulation of genetic alterations in thyroid follicular cells, creating a favorable environment for the initiation and progression of FTC. Aberrant BMP signaling supports tumor growth by enhancing cell proliferation, angiogenesis, and resistance to apoptosis. FTC often arises from thyroid follicular cells that have undergone oncogenic transformation due to sustained disruption of BMP signaling pathways by NF-κB [103, 104].

#### 13. Notch signaling pathway Role of NF-κB in disrupting it

Notch signaling plays a critical role in thyroid development, influencing cell differentiation, proliferation, and tissue patterning. It regulates the differentiation of thyroid progenitor cells into functional thyroid follicular cells and is involved in maintaining proper tissue morphogenesis and growth during thyroid gland development. Dysregulation of Notch signaling due to inflammation, including NF-κB activation, can significantly impact thyroid function and contribute to thyroid disorders. Inflammatory cytokines such as TNF-α and IL-1β induce NF-κB activation in response to thyroid inflammation [105]. NF-κB then translocates to the nucleus, where it regulates the expression of genes involved in inflammation and cellular responses, which can potentially affect Notch signaling components. Inflammatory conditions also induce oxidative stress in thyroid cells, which can further modulate Notch signaling pathways involved in cell fate determination and proliferation. NF-κB activation can influence the expression levels of Notch receptors, such as Notch1-4, and ligands, including Jagged and Delta-like, on thyroid follicular cells. Altered expression of these Notch components can disrupt the balance of Notch signaling, affecting downstream events essential for thyroid cell differentiation and function [106]. Additionally, NF-κB-mediated inflammation can interact with intracellular signaling pathways activated by Notch, such as the NICD-RBPJκ complex, leading to aberrant activation of downstream effectors. This dysregulation can influence gene expression profiles and cellular responses in thyroid follicular cells. Dysregulated Notch signaling under inflammatory conditions can result in abnormal thyroid gland development, which may manifest as either reduced or excessive tissue growth, known as hypoplasia or hyperplasia. Aberrant Notch signaling is also associated with thyroid cancer development, including papillary thyroid carcinoma (PTC) and possibly other types through interactions with other pathways [107]. Furthermore, altered Notch signaling can disrupt thyroid hormone synthesis and secretion, contributing to hypothyroidism or thyroid hormone resistance. Notch signaling pathway activation in thyroid tumors may serve as a prognostic indicator for disease aggressiveness and response to targeted therapies. NF-κB activation disrupts Notch signaling pathways in thyroid follicular cells, altering cell fate regulation and increasing susceptibility to various thyroid disorders, including cancer. Dysregulated Notch signaling under inflammatory conditions promotes abnormal cell proliferation, survival, and differentiation, which contributes to thyroid dysfunction and disease progression. Further research into the interplay between NF-κB and Notch signaling pathways is essential for developing effective therapeutic strategies and improving clinical outcomes in patients with thyroid diseases [108].

##### Resultant dysregulation in thyroid cell fate and resultant impact on FTC development

NF-κB activation can significantly impact Notch signaling in thyroid follicular cells, potentially disrupting cell fate regulation and increasing the risk of follicular thyroid carcinoma (FTC). Inflammatory cytokines such as TNF-α and IL-1β trigger NF-κB activation in response to thyroid inflammation. Once activated, NF-κB translocates to the nucleus where it regulates the expression of genes involved in inflammation and cellular responses, which can affect Notch signaling components. This activation can also influence the expression levels of Notch receptors, including Notch1-4, and ligands such as Jagged and Delta-like, on thyroid follicular cells. Altered Notch expression levels can disrupt the balance of Notch signaling, impacting downstream signaling events important for thyroid cell differentiation and function [109]. Furthermore, NF-κB-mediated inflammation can interact with intracellular signaling pathways activated by Notch, such as the NICD-RBPJκ complex. This interaction can lead to aberrant activation of downstream effectors, influencing gene expression profiles and cellular responses in thyroid follicular cells. Dysregulated Notch signaling under the influence of NF-κB can disrupt normal differentiation programs, potentially resulting in a dedifferentiated state where thyroid cells lose their specialized functions and acquire stem-like properties, which may promote oncogenic transformation [110]. Additionally, NF-κB-induced Notch signaling can enhance thyroid cell proliferation and survival mechanisms, creating a cellular environment conducive to oncogenic transformation, particularly towards follicular thyroid carcinoma. Dysregulated Notch signaling, influenced by NF-κB activation, promotes genomic instability and accumulation of genetic alterations in thyroid follicular cells, creating a favorable environment for the initiation and progression of follicular thyroid carcinoma. This aberrant Notch signaling supports tumor growth by enhancing cell proliferation, angiogenesis, and resistance to apoptosis, leading to FTC from thyroid follicular cells that have undergone oncogenic transformation due to sustained NF-κB-mediated disruption of Notch signaling pathways [111, 112].

#### 14. TGF-β

##### Role of NF-κB in disrupting it

Transforming Growth Factor-beta (TGF-β) signaling plays a key role in thyroid development by influencing cell differentiation, proliferation, and tissue remodeling. Dysregulation of TGF-β signaling due to inflammation, including NF-κB activation, can impact thyroid function and contribute to thyroid disorders. TGF-β signaling is essential for regulating the differentiation of thyroid progenitor cells into functional thyroid follicular cells and plays a role in maintaining proper tissue morphogenesis and growth during thyroid gland development [113]. Inflammatory cytokines such as TNF-α and IL-1β induce NF-κB activation in response to thyroid inflammation. Activated NF-κB translocates to the nucleus, where it regulates the expression of genes involved in inflammation and cellular responses, potentially affecting TGF-β signaling components. Inflammatory conditions also induce oxidative stress in thyroid cells, which can modulate TGF-β signaling pathways involved in cell fate determination and proliferation. NF-κB activation can influence the expression levels of TGF-β ligands, such as TGF-β1, TGF-β2, and TGF-β3, as well as their receptors, including TGF-β receptor types I, II, and III, on thyroid follicular cells. Altered TGF-β expression levels can disrupt the balance of TGF-β signaling, affecting downstream signaling events critical for thyroid cell differentiation and function [114]. Moreover, NF-κB-mediated inflammation can interact with intracellular signaling pathways activated by TGF-β, such as SMAD-dependent and non-SMAD pathways. Dysregulated TGF-β signaling can result in aberrant activation of downstream effectors, influencing gene expression profiles and cellular responses in thyroid follicular cells. The consequences of dysregulated TGF-β signaling under inflammatory conditions include abnormal thyroid gland development, manifesting as either reduced or excessive tissue growth, known as hypoplasia or hyperplasia. Altered TGF-β signaling is also associated with thyroid cancer development, including follicular and papillary thyroid carcinoma, through its effects on cell proliferation, invasion, and metastasis. Additionally, dysregulated TGF-β signaling disrupts thyroid hormone synthesis and secretion, contributing to hypothyroidism or thyroid hormone resistance. Assessment of TGF-β signaling pathway activation in thyroid tumors may serve as a prognostic indicator for disease aggressiveness and response to targeted therapies. NF-κB activation disrupts TGF-β signaling pathways in thyroid follicular cells, affecting cell fate regulation and predisposing the thyroid gland to various disorders, including cancer. Dysregulated TGF-β signaling under inflammatory conditions promotes abnormal cell proliferation, survival, and differentiation, contributing to thyroid dysfunction and disease progression. Further research into the interplay between NF-κB and TGF-β signaling pathways is essential for developing effective therapeutic strategies and improving clinical outcomes in patients with thyroid diseases [115, 116].

##### Resultant dysregulation in thyroid cell fate and resultant impact on FTC development

NF-κB activation can significantly impact Transforming Growth Factor-beta (TGF-β) signaling in thyroid follicular cells, potentially disrupting cell fate regulation and contributing to the predisposition of the thyroid gland to follicular thyroid carcinoma (FTC). Inflammatory cytokines such as TNF-α and IL-1β induce NF-κB activation in response to thyroid inflammation. Once activated, NF-κB translocates to the nucleus and regulates the expression of genes involved in inflammation and cellular responses, which can potentially affect TGF-β signaling components [117]. NF-κB activation can also influence the expression levels of TGF-β ligands, including TGF-β1, TGF-β2, and TGF-β3, as well as their receptors, such as TGF-β receptor types I, II, and III, on thyroid follicular cells. Altered TGF-β expression levels can disrupt the balance of TGF-β signaling, affecting downstream signaling events that are critical for thyroid cell differentiation and function. Additionally, NF-κB-mediated inflammation can interact with intracellular signaling pathways activated by TGF-β, including both SMAD-dependent and non-SMAD pathways. This dysregulation can lead to aberrant activation of downstream effectors, influencing gene expression profiles and cellular responses in thyroid follicular cells. Dysregulated TGF-β signaling due to NF-κB influence can disrupt the normal differentiation programs of thyroid follicular cells, potentially leading to a dedifferentiated state where thyroid cells lose their specialized functions and acquire stem-like properties conducive to oncogenic transformation [118]. The activation of TGF-β signaling by NF-κB can also enhance thyroid cell proliferation and survival mechanisms. This increased cell proliferation, combined with impaired differentiation, creates a cellular environment that is prone to oncogenic transformation, particularly towards follicular thyroid carcinoma. Dysregulated TGF-β signaling, influenced by NF-κB activation, promotes genomic instability and the accumulation of genetic alterations in thyroid follicular cells, creating a favorable environment for the initiation and progression of follicular thyroid carcinoma. Aberrant TGF-β signaling under the influence of NF-κB supports tumor growth by enhancing cell proliferation, angiogenesis, and resistance to apoptosis, leading to FTC arising from thyroid follicular cells that have undergone oncogenic transformation due to sustained NF-κB-mediated disruption of TGF-β signaling pathways [119, 120].

## Discussion

### Landscape of Dysregulations in Thyroid Genes/TFs/Pathways Induced by NF-κB Activation

#### 1. Genes/Transcription Factors

- NKX2-1

- **Normal Function**: Essential for thyroid follicular cell differentiation and function.
- **NF-κB Impact**: Downregulation leads to loss of differentiation and increased cellular proliferation, promoting a dedifferentiated, oncogenic state.
- PAX8

- **Normal Function**: Critical for thyroid follicular cell development and hormone synthesis.
- **NF-κB Impact**: Suppression results in disrupted cellular identity and increased susceptibility to malignancy.
- FOXE1 (TTF-2)

- **Normal Function**: Involved in thyroid gland morphogenesis and differentiation.
- **NF-κB Impact**: Reduced expression compromises differentiation and function, creating an environment favorable for tumorigenesis.
- HHEX

- **Normal Function**: Regulates early thyroid development and cell differentiation.
- **NF-κB Impact**: Dysregulation impairs normal development and promotes aberrant cell growth.

#### 2. Signaling Pathways

- TSH Signaling Pathway

- **Normal Function**: Regulates thyroid hormone synthesis and secretion.
- **NF-κB Impact**: Disruption alters hormone production, contributing to dedifferentiation and tumorigenesis.
- RET/PTC

- **Normal Function**: Involved in thyroid cell growth and differentiation.
- **NF-κB Impact**: Aberrant signaling enhances proliferative and anti-apoptotic pathways, supporting tumor growth.
- MAPK/ERK Pathway

- **Normal Function**: Controls cell proliferation, differentiation, and survival.
- **NF-κB Impact**: Dysregulation promotes uncontrolled cell proliferation and survival, increasing oncogenic potential.
- PI3K/AKT Pathway

- **Normal Function**: Regulates cell growth, proliferation, and survival.
- **NF-κB Impact**: Enhanced pathway activity leads to increased cell proliferation and survival, predisposing to malignancy.
- Wnt/β-catenin Pathway

- **Normal Function**: Governs cell proliferation, differentiation, and tissue homeostasis.
- **NF-κB Impact**: Dysregulation disrupts differentiation and promotes tumorigenesis.
- FGF Signaling

- **Normal Function**: Regulates cell proliferation, differentiation, and survival.
- **NF-κB Impact**: Altered signaling affects growth and differentiation processes, increasing cancer risk.
- Shh Pathway

- **Normal Function**: Involved in tissue patterning and cell differentiation.
- **NF-κB Impact**: Disruption affects proliferation and differentiation, contributing to oncogenic transformation.
- BMP Signaling

- **Normal Function**: Controls cell growth and differentiation.
- **NF-κB Impact**: Dysregulation alters growth and differentiation, promoting malignancy.
- Notch Pathway

- **Normal Function**: Regulates cell differentiation and proliferation.
- **NF-κB Impact**: Modulation impairs differentiation and promotes a stem-like, oncogenic phenotype.
- TGF-β Signaling

- **Normal Function**: Regulates cell growth, differentiation, and apoptosis.
- **NF-κB Impact**: Dysregulation promotes aberrant cell proliferation and survival, fostering a tumorigenic environment.

### Pathological State Induction

The cumulative effect of NF-κB activation and resultant dysregulation of these genes, transcription factors and signaling pathways leads to:

- **Loss of Differentiation**: Cells lose their specialized functions and acquire a dedifferentiated, stem-like state.
- **Increased Proliferation**: Enhanced cell division and survival mechanisms.
- **Genomic Instability**: Increased susceptibility to genetic mutations and chromosomal aberrations.
- **Oncogenic Transformation**: Conditions favorable for the initiation and progression of follicular thyroid carcinoma (FTC).

Chronic inflammation and NF-κB activation result in widespread dysregulation of transcription factors and signaling pathways essential for thyroid development and function. This dysregulation leads to a loss of thyroid follicular cell fate integrity, increased proliferation, and genomic instability, ultimately predisposing the thyroid gland to follicular thyroid carcinoma.

### Net Effect

- **Proliferation Increase**: NF-κB activation generally leads to upregulation of pathways that promote cell proliferation (e.g., MAPK/ERK, PI3K/AKT), contributing to increased cell growth and survival.
- **Differentiation Decrease**: Concurrently, NF-κB suppresses the expression and function of transcription factors and pathways critical for thyroid follicular cell differentiation (e.g., NKX2-1, PAX8, FOXE1, TSH signaling), leading to impaired maturation of thyroid cells.

NF-κB-induced damage in thyroid follicular cells disrupts the delicate balance between proliferation and differentiation. By downregulating key transcription factors and dysregulating essential signaling pathways, NF-κB promotes a proliferative, undifferentiated state in thyroid cells. This dysregulation not only impairs normal thyroid function but also predisposes the gland to oncogenic transformation, particularly follicular thyroid carcinoma.

### Clinical Implications of NF-κB Induced Dysregulations in Thyroid Follicular Cells

Understanding the impact of NF-κB on key transcription factors such as NKX2-1, PAX8, FOXE1, and HHEX, as well as on critical signaling pathways including TSH, RET/PTC, MAPK/ERK, PI3K/AKT, Wnt/β-catenin, FGF, Shh, BMP, Notch, and TGF-β, is essential for identifying biomarkers for the early detection of thyroid dysregulation and follicular thyroid carcinoma (FTC). Implementing molecular profiling in patients with chronic thyroid inflammation can aid in detecting early signs of NF-κB activation and subsequent disruptions in thyroid function, facilitating early diagnosis of FTC. The development of targeted therapies aimed at inhibiting NF-κB activation or restoring the normal function of affected transcription factors and signaling pathways could potentially prevent the progression from chronic inflammation to FTC. Combining NF-κB inhibitors with therapies that target specific disrupted pathways, such as MAPK/ERK and PI3K/AKT, could enhance therapeutic efficacy and prevent resistance.

Assessing the status of NF-κB activation and its downstream effects on thyroid-specific genes and pathways can help stratify patients based on their risk of developing FTC, informing clinical decisions regarding surveillance and intervention strategies. Evaluating the extent of NF-κB-induced dysregulation in thyroid follicular cells could provide valuable prognostic information, assisting in predicting disease progression and patient outcomes. Implementing anti-inflammatory interventions in patients with thyroid conditions can help mitigate NF-κB activation, potentially preventing the dysregulation of thyroid genes and pathways and reducing the risk of FTC. Encouraging lifestyle and dietary modifications that reduce inflammation may serve as preventive measures against NF-κB activation and its detrimental effects on thyroid health. Personalized treatment plans based on the molecular profile of NF-κB activation and its impact on thyroid-specific factors can improve patient outcomes. Tailoring therapies to address specific dysregulations can enhance treatment effectiveness and reduce side effects. Additionally, genetic screening for mutations in NF-κB-related pathways and thyroid-specific genes may identify individuals at higher risk for developing FTC, allowing for personalized surveillance and early intervention. Insights into the mechanisms by which NF-κB disrupts thyroid cell proliferation and differentiation provide a foundation for developing possible new drugs targeting these pathways. Research efforts can focus on creating inhibitors that specifically block NF-κB or its downstream effects. Conducting clinical trials to test the efficacy of NF-κB inhibitors and combination therapies in patients with thyroid inflammation and early-stage FTC can validate their clinical utility and establish new treatment protocols. The findings from this research highlight the key role of NF-κB in disrupting thyroid follicular cell proliferation and differentiation, predisposing the thyroid gland to follicular thyroid carcinoma. Translating these insights into clinical practice can improve diagnostic, therapeutic, and prognostic approaches, ultimately enhancing patient care and outcomes in thyroid diseases. By targeting the underlying molecular mechanisms, more effective and personalized strategies can be developed to prevent and treat FTC, addressing a significant clinical challenge in thyroid pathology.

### Key Findings

1. NF-κB Activation and NKX2-1 Suppression:

- **Finding**: Chronic NF-κB activation leads to the suppression of NKX2-1, a key transcription factor essential for thyroid follicular cell differentiation and function.
- **Implication**: This suppression results in the dedifferentiation of thyroid cells, predisposing them to oncogenic transformation and increasing the risk of FTC.
2. PAX8 Dysregulation:

- **Finding**: NF-κB-induced inflammation disrupts the normal expression of PAX8, a critical transcription factor in thyroid development and function.
- **Implication**: This dysregulation impairs thyroid cell differentiation and promotes a cellular environment conducive to malignant transformation.
3. FOXE1 (TTF-2) Inhibition:

- **Finding**: Inflammatory signaling via NF-κB inhibits FOXE1, another key transcription factor for thyroid development.
- **Implication**: Reduced FOXE1 activity leads to disrupted thyroid morphogenesis and function, contributing to increased FTC risk.
4. HHEX Disruption:

- **Finding**: NF-κB activation interferes with HHEX, critical for early thyroid development.
- **Implication**: This disruption may lead to aberrant thyroid tissue architecture and function, fostering conditions favorable for FTC development.
5. TSH Signaling Pathway Impairment:

- **Finding**: NF-κB negatively impacts the TSH signaling pathway, vital for thyroid hormone production and cell
- proliferation.
- **Implication**: Impaired TSH signaling results in altered thyroid homeostasis and increased susceptibility to FTC.
6. RET/PTC Rearrangement Enhancement:

- **Finding**: NF-κB activity promotes RET/PTC rearrangements, known oncogenic drivers in thyroid cancer.
- **Implication**: Enhanced RET/PTC rearrangements contribute to the malignant transformation of thyroid follicular cells.
7. MAPK/ERK Pathway Alteration:

- **Finding**: NF-κB modifies the MAPK/ERK pathway, essential for cell growth and differentiation.
- **Implication**: Alterations in this pathway lead to uncontrolled cell proliferation and increased FTC risk.
8. PI3K/AKT Pathway Activation:

- **Finding**: Chronic inflammation mediated by NF-κB activates the PI3K/AKT pathway, associated with cell survival and growth.
- **Implication**: This activation supports the survival of aberrant cells, promoting FTC development.
9. Wnt/β-catenin Pathway Dysregulation:

- **Finding**: NF-κB disrupts the Wnt/β-catenin pathway, essential for thyroid cell proliferation and differentiation.
- **Implication**: Dysregulation of this pathway contributes to thyroid follicular cell dedifferentiation and oncogenesis.
10. FGF Signaling Interference:

- **Finding**: Inflammatory signals via NF-κB interfere with FGF signaling, important for thyroid development.
- **Implication**: This interference impairs thyroid tissue growth and repair, facilitating conditions for FTC.
11. Shh Pathway Modulation:

- **Finding**: NF-κB modulates the Shh pathway, involved in organogenesis and tissue patterning.
- **Implication**: Modulation of Shh by NF-κB disrupts thyroid gland development, increasing FTC susceptibility.
12. BMP Pathway Disruption:

- **Finding**: NF-κB disrupts BMP signaling, which plays a role in thyroid follicular cell differentiation.
- **Implication**: Disruption in BMP signaling leads to abnormal thyroid development and predisposes cells to malignancy.
13. Notch Pathway Alteration:

- **Finding**: NF-κB alters Notch signaling, critical for maintaining thyroid cell fate.
- **Implication**: Altered Notch signaling results in disrupted thyroid cell differentiation, increasing the risk of FTC.
14. TGF-β Pathway Dysregulation:

- **Finding**: NF-κB dysregulates the TGF-β pathway, involved in cellular proliferation and differentiation.
- **Implication**: Dysregulation of TGF-β signaling promotes a pro-tumorigenic environment in the thyroid gland.

## Conclusion

Chronic inflammation and resultant NF-κB activation disrupt the regulatory networks governing thyroid follicular cell fate, ultimately predisposing the thyroid gland to follicular thyroid carcinoma (FTC). NF-κB-mediated inflammation leads to the downregulation of NKX2-1, impairing thyroid follicular cell differentiation and promoting a more proliferative, undifferentiated cell state. Inflammatory conditions and NF-κB activation also result in the suppression of PAX8, which disrupts thyroid follicular cell development and function, leading to a loss of cellular identity and enhanced susceptibility to malignancy. The impact of NF-κB extends to FOXE1 (TTF-2), where its expression is compromised, further impairing differentiation and proper functioning of thyroid follicular cells, thereby facilitating a cellular environment conducive to oncogenic transformation. Additionally, dysregulation of HHEX due to NF-κB activation hampers its role in thyroid development, contributing to aberrant cell growth and differentiation. NF-κB disrupts the TSH signaling pathway, leading to altered thyroid hormone synthesis and secretion, which may contribute to cellular dedifferentiation and tumorigenesis. It also influences RET/PTC signaling, promoting proliferative and anti-apoptotic pathways, thereby enhancing tumor growth and progression. Activation of NF-κB results in the aberrant regulation of the MAPK/ERK pathway, fostering uncontrolled cell proliferation and survival. NF-κB-mediated dysregulation of the PI3K/AKT pathway enhances thyroid follicular cell proliferation and survival, predisposing cells to oncogenic changes. Inflammatory conditions activate NF-κB, leading to dysregulation of Wnt/β-catenin signaling, which disrupts cellular differentiation and promotes tumorigenesis. NF-κB also influences FGF signaling, altering cell proliferation and differentiation processes critical for maintaining thyroid follicular cell identity. Furthermore, NF-κB-mediated inflammation disrupts Shh signaling, affecting cell proliferation and differentiation during thyroid development and contributing to malignancy. The impact of NF-κB on BMP signaling leads to alterations in thyroid follicular cell growth and differentiation, thereby increasing the risk of cancer development. Additionally, NF-κB activation modulates Notch signaling, impairing thyroid follicular cell differentiation and promoting a stem-like, oncogenic phenotype. Dysregulation of TGF-β signaling by NF-κB disrupts thyroid cell differentiation and promotes a tumorigenic environment. The cumulative effect of NF-κB activation and the resultant dysregulation of these critical factors and pathways creates a cellular milieu characterized by reduced differentiation, increased proliferation, and enhanced survival. This aberrant state fosters genomic instability and the accumulation of oncogenic mutations, predisposing thyroid follicular cells to transformation into FTC. The interplay between chronic inflammation, NF-κB activation, and disrupted thyroid signaling networks points towards the importance of targeted therapeutic strategies to restore normal signaling and prevent malignant transformation in thyroid diseases. By delineating these mechanisms, our findings provide a comprehensive understanding of how chronic inflammation and NF-κB activation contribute to thyroid carcinogenesis, offering potential avenues for the development of targeted treatments and improved prognostic markers for FTC.

## Abbreviations

NF-κB: Nuclear Factor Kappa-Light-Chain-Enhancer of Activated B Cells
NKX2-1: NK2 Homeobox 1
PAX8: Paired Box 8
FOXE1: Forkhead Box E1 (Thyroid Transcription Factor 2, TTF-2)
HHEX: Hematopoietically Expressed Homeobox
TSH: Thyroid Stimulating Hormone
RET/PTC: Rearranged during Transfection/Papillary Thyroid Carcinoma
MAPK/ERK: Mitogen-Activated Protein Kinase/Extracellular Signal-Regulated Kinase
PI3K/AKT: Phosphoinositide 3-Kinase/Protein Kinase B
Wnt/β-catenin: Wnt Signaling Pathway/Beta-Catenin
FGF: Fibroblast Growth Factor
Shh: Sonic Hedgehog
BMP: Bone Morphogenetic Protein
Notch: Notch Signaling Pathway
TGF beta: Transforming Growth Factor Beta

## Declarations

### Ethics declarations

Ethics approval and consent to participate

Not applicable.

### Consent for publication

Not applicable.

### Data Availability statement

All data generated or analyzed during this study are included in this article.

### Competing interests

The authors declare that they have no competing interests.

### Funding

I declare that there was not any source of funding for this research work.

### Acknowledgements

“Not applicable”.

### Authors’ Information

**1. Ovais Shafi (OS)\*** is the author of the study and was involved in the idea, concept, design, and methodology of the study, literature search and references. He did the writing, editing, and revision of the manuscript. He was involved in drawing the findings, results, conclusions, implications of the study, interpretation of the data and was involved in all aspects of the study. He prepared and wrote discussion, results, conclusions and all areas of the study. OS extracted and analyzed the data. He was involved in critical evaluation, audit of every aspect of the study, data extraction, adherence of the study to relevant PRISMA guidelines, limitations of the study, references, and all others. He was involved in drawing PRISMA Flow Diagram. The author read and approved the manuscript.

He investigated the thyroid genes/tfs/signaling pathways for their roles in relation to this study:

NKX2-1, PAX8, FOXE1 (TTF-2), HHEX, TSH, RET/PTC, MAPK/ERK Pathway, PI3K/AKT Pathway, Wnt/β-catenin Pathway, FGF, Shh, BMP, Notch, TGF beta.

**Ovais Shafi (OS)\***, MBBS - Sindh Medical College - Dow University of Health Sciences, Karachi, Pakistan. He aspires to become an eminent ‘Physician Scientist’. He is devoted to the research in disease development mechanisms, disease origins and therapeutics. OS is also passionate about multiple research areas including clinical trials, clinical medicine, therapeutics, regenerative medicine, precision medicine including gene therapies, finding disease specific targets for gene therapy, role of disease genomics and epigenetics in diagnosis, management, and therapeutics development. He is dedicated to the field of research and clinical medicine. Email address*

Corresponding author: OS

Correspondence to Ovais Shafi

2. **Raveena (RA)** is the co-author of the study. She contributed to the results and conclusions of the study, also contributed to the writing and editing of these sections. She contributed to investigating the thyroid genes/tfs/signaling pathways for their roles in relation to this study:

NKX2-1, PAX8, FOXE1 (TTF-2), HHEX, TSH, RET/PTC, MAPK/ERK Pathway, PI3K/AKT Pathway, Wnt/β-catenin Pathway, FGF, Shh, BMP, Notch, TGF beta.

Raveena, MBBS - Sindh Medical College – Jinnah Sindh Medical University, Karachi, Pakistan. She is passionate about research in surgery and disease development mechanisms including neurodegenerative diseases, oncogenesis and others. She is ECFMG Certified. She is passionate about residency in Internal Medicine/Surgery. Her goal is to make significant impact in the field of Research.

3. **Fatima Arshad (FA)** is the co-author of the study. She contributed to the results and conclusions of the study, also contributed to the writing and editing of these sections. She contributed to investigating the thyroid genes/tfs/signaling pathways for their roles in relation to this study:

NKX2-1, PAX8, FOXE1 (TTF-2), HHEX, TSH, RET/PTC, MAPK/ERK Pathway, PI3K/AKT Pathway, Wnt/β-catenin Pathway, FGF, Shh, BMP, Notch, TGF beta.

Fatima Arshad, MBBS - Sindh Medical College - Dow University of Health Sciences, Karachi, Pakistan. She is passionate about research in disease development mechanisms with potential implications towards better patient outcomes.

4. **Finza Kanwal (FK)** is the co-author of the study. She contributed to the results and conclusions of the study, also contributed to the writing and editing of these sections. She contributed to investigating the thyroid genes/tfs/signaling pathways for their roles in relation to this study:

NKX2-1, PAX8, FOXE1 (TTF-2), HHEX, TSH, RET/PTC, MAPK/ERK Pathway, PI3K/AKT Pathway, Wnt/β-catenin Pathway, FGF, Shh, BMP, Notch, TGF beta.

Finza Kanwal, MBBS - Sindh Medical College - Dow University of Health Sciences, Karachi, Pakistan. She has been affiliated with Liaquat National Hospital and Medical College, Pakistan. Currently, she is affiliated with The College of Physicians and Surgeons Pakistan. Her goal is to be an Endocrinologist (MRCP).

5. **Osama Jawed Khan (OJK)** is the co-author of the study. He contributed to the results and conclusions of the study, also contributed to the writing and editing of these sections. He contributed to investigating the thyroid genes/tfs/signaling pathways for their roles in relation to this study:

NKX2-1, PAX8, FOXE1 (TTF-2), HHEX, TSH, RET/PTC, MAPK/ERK Pathway, PI3K/AKT Pathway, Wnt/β-catenin Pathway, FGF, Shh, BMP, Notch, TGF beta.

Osama Jawed Khan, MBBS - Sindh Medical College - Dow University of Health Sciences, Karachi, Pakistan. He has been affiliated with Dr. Ruth K. M. Pfau, Civil Hospital Karachi, Pakistan. Currently, he is affiliated with Royal Institute of Medicine & Surgery (RIMS), Karachi. He intends to pursue fellowship of the Royal Colleges of Surgeons (FRCS).

6. **Muhammad Ashar (MA)** is the co-author of the study. He contributed to the results and conclusions of the study, also contributed to the writing and editing of these sections. He contributed to investigating the thyroid genes/tfs/signaling pathways for their roles in relation to this study:

NKX2-1, PAX8, FOXE1 (TTF-2), HHEX, TSH, RET/PTC, MAPK/ERK Pathway, PI3K/AKT Pathway, Wnt/β-catenin Pathway, FGF, Shh, BMP, Notch, TGF beta.

Muhammad Ashar, MBBS/MD is a Resident Medical Officer in the Department of General Surgery at Aga Khan University Hospital, Karachi, Pakistan. He has extensive clinical experience across Pakistan and the United States. His research focuses on hepatocellular carcinoma and other gastrointestinal malignancies. He is ECFMG certified and holds a medical license in the United States, with plans to pursue further training and a career in oncology in the US. He is affiliated with Aga Khan University Hospital, Department of General Surgery, Hepato-Pancreato-Biliary and Surgical Oncology, Karachi, Pakistan. His research interests include Hepato-pancreato-biliary cancers, Gastrointestinal oncology, Esophageal cancer.

7. **Abdul Jabbar (AJ)** is the co-author of the study. He contributed to the results and conclusions of the study, also contributed to the writing and editing of these sections. He contributed to investigating the thyroid genes/tfs/signaling pathways for their roles in relation to this study:

NKX2-1, PAX8, FOXE1 (TTF-2), HHEX, TSH, RET/PTC, MAPK/ERK Pathway, PI3K/AKT Pathway, Wnt/β-catenin Pathway, FGF, Shh, BMP, Notch, TGF beta.

Abdul Jabbar, MBBS - Sindh Medical College - Jinnah Sindh Medical University, Karachi, Pakistan. He is currently a Medical officer in an NGO. He is interested in research in disease development mechanisms. He is ECFMG Certified, his goal is to do Medical residency in Internal Medicine. He aspires to be a specialist in Cardiology.

7. **Haresh Kumar (HK)** is the co-author of the study. He contributed to the results and conclusions of the study, also contributed to the writing and editing of these sections. He contributed to investigating the thyroid genes/tfs/signaling pathways for their roles in relation to this study:

NKX2-1, PAX8, FOXE1 (TTF-2), HHEX, TSH, RET/PTC, MAPK/ERK Pathway, PI3K/AKT Pathway, Wnt/β-catenin Pathway, FGF, Shh, BMP, Notch, TGF beta.

Haresh Kumar, MBBS, Diploma in Diabetes and Endocrinology, Plab 1 & 2 MRCP UK 1. He completed his med-school from Shaheed Mohtarma Benazir Bhutto Medical College, Sindh Government Lyari General Hospital. His future goals include pursuing career with NHS, UK.

8. **Rahimeen Rajpar (RR)** is also the author of the study and contributed to the results and conclusions of this study along with working on the findings. She contributed to investigating the thyroid genes/tfs/signaling pathways for their roles in relation to this study:

NKX2-1, PAX8, FOXE1 (TTF-2), HHEX, TSH, RET/PTC, MAPK/ERK Pathway, PI3K/AKT Pathway, Wnt/β-catenin Pathway, FGF, Shh, BMP, Notch, TGF beta.

Rahimeen Rajpar, MD is a dedicated medical professional with a passion for unraveling the mysteries of disease origins and progression. Currently pursuing her residency in internal medicine. RR is committed to advancing the field of medical research. Her goal is to become a leader in the field of Medicine, looking at the medical intricacies from a different lens. Apart from research RR remains dedicated to improving the lives of her patients through comprehensive care and by working towards scientific discoveries.

9. **Khizer Sohail (KS)** is the co-author of the study. He contributed to the results and conclusions of the study, also contributed to the writing and editing of these sections. He contributed to investigating the thyroid genes/tfs/signaling pathways for their roles in relation to this study:

NKX2-1, PAX8, FOXE1 (TTF-2), HHEX, TSH, RET/PTC, MAPK/ERK Pathway, PI3K/AKT Pathway, Wnt/β-catenin Pathway, FGF, Shh, BMP, Notch, TGF beta.

Khizer Sohail, MBBS, from Jinnah Sindh Medical University in Karachi, Pakistan, is a committed medical professional with a solid foundation in clinical medicine and healthcare. His academic journey has sparked a deep interest in oncology, focusing on advancing the understanding and treatment of cancer. He is particularly passionate about researching the origins and development mechanisms of diseases, with a specific interest in the molecular and cellular processes underlying tumour formation. Additionally, he is dedicated to research improving management strategies and patient outcomes in toxicology. Driven by a desire to advance medical science, he aims to pursue a residency in Internal Medicine. He hopes to integrate research insights towards clinical practice to innovate and enhance health outcomes.

10. **Manwar Madhwani (MM)** is the co-author of the study. He contributed to the results of the study. He contributed to investigating the thyroid genes/tfs/signaling pathways for their roles in relation to this study:

NKX2-1, PAX8, FOXE1 (TTF-2), HHEX, TSH, RET/PTC, MAPK/ERK Pathway, PI3K/AKT Pathway, Wnt/β-catenin Pathway, FGF, Shh, BMP, Notch, TGF beta.

Manwar Madhwani, MBBS - Sindh Medical College – Jinnah Sindh Medical University, Karachi, Pakistan. He is currently a medical officer in an NGO. He is interested in research in disease development mechanisms. His goal is to do Medical Residency in Internal Medicine after completing his ECFMG Certification. He aspires to be a specialist in Cardiology/ Pediatrics.

11. **Muhammad Waqas (MW)** is the co-author of the study. He contributed to the results of the study. He contributed to investigating the thyroid genes/tfs/signaling pathways for their roles in relation to this study:

NKX2-1, PAX8, FOXE1 (TTF-2), HHEX, TSH, RET/PTC, MAPK/ERK Pathway, PI3K/AKT Pathway, Wnt/β-catenin Pathway, FGF, Shh, BMP, Notch, TGF beta.

Muhammad Waqas, MBBS - Sindh Medical College - Dow University of Health Sciences, Karachi, Pakistan. He is ECFMG Certified. His future goals include residency in Internal Medicine/Neurology, and fellowship in Cardiology/Nephrology/Critical Care.

*The work and contributions of everyone have been described in detail, the order is randomized and the numbering is just for referencing purpose.*

## Notes

### Competing Interest Statement

The authors have declared no competing interest.

